# Conversion to Parkinsonism and Dementia in REM-Sleep Behavior Disorder Using the Chronobiotic Melatonin

**DOI:** 10.1101/2020.11.05.20224592

**Authors:** Dieter Kunz, Sophia Stotz, Frederik Bes

## Abstract

**Background:** Isolated REM sleep behavior disorder (iRBD), a reliable prodromal stage marker of α-synucleinopathies like Parkinson’s disease or Lewy body dementia, offers an early window for disease-modifying intervention. Current treatments of iRBD, including the two level B therapies with clonazepam and melatonin, are considered symptomatic. However, numbers of reported patients treated with melatonin are low and whether melatonin has disease-modifying potential is unclear.

**Methods:** This single-center, prospective cohort study included 206 consecutive patients diagnosed with iRBD until January 2020. Thirty-nine patients had applied mixed treatments on the advice of the referring physician, 167 had administered melatonin according to our chronobiotic protocol (low dose, ≥ 6 months, *always-at-the-same-clock-time*, between 10 and 11 pm - corrected for chronotype), which differs from existing melatonin prescriptions. Clinical examination to determine phenoconversion was performed from October 2018 to August 2020. To evaluate generalizability, we compared factors such as neuropsychological and neuromotor performance, olfactory ability, neurovegetative behavior, and dopamine transporter density in our patients with those reported for other cohorts. Primary outcome was phenoconversion to clinical synucleinopathy, assessed using Kaplan-Meier analysis. Secondary outcomes were changes in cognitive and motor performance, and in RBD-symptom severity, analyzed using mixed models.

**Results:** RBD characteristics were comparable to those in other published cohorts, including frequency of phenoconversion in our patients with mixed treatments (10/39; follow-up 3.1±2.1 years). In contrast, long-term melatonin-treated patients rarely converted (4/167; follow-up 4.2±3.1 years; hazard-ratio 0.07, 95% CI, 0.02-0.22, p<0.001). Neuromotor and neuropsychological performance did not decline, improved in some domains. Symptom severity gradually improved over the first 4 weeks of treatment (Clinical Global Impression Severity: 5.7 vs. 3.0) and remained stable over years, also in those patients who had stopped melatonin intake after 6 months. The initial response was slower in patients with melatonin suppressing (beta blockers) or REM sleep spoiling co-medication (antidepressants) and failed with inadequate timing of melatonin intake.

**Conclusion:** Clock-timed melatonin treatment in patients with iRBD appears to be associated with a marked reduction in the development of parkinsonism and dementia as well as with an improvement in neuromotor, cognitive, and specific RBD symptoms. Findings suggest that melatonin treatment may have disease-modifying effects in synucleinopathies. The fact that melatonin is available anywhere at low cost provides the perspective of immediate clinical application in patients at risk for clinical synucleinopathy. On the other hand, clock-time dependency challenges existing prescription guidelines for melatonin. Melatonin should be acknowledged as the darkness signal to circadian clock-work rather than a hypnotic.

## INTRODUCTION

Parkinsonism and dementia caused by neurodegeneration are characterized by gradual, irreversible worsening of physical and mental abilities, leading to patient distress and dysautonomy, severe burden for care givers, and high socio-economic costs (GBD, 2016). Knowledge of the disease has improved substantially in recent years, and effective symptomatic treatment has been identified, improving patients’ quality of life. Attempts to modify the disease, however, have not been successful to date (Espay *et al*., 2017, 2020). The clinical signs of parkinsonism and dementia occur when the underlying neurodegenerative process is already far advanced (Braak *et al*., 2003). Hence, a major challenge of research in this field is to identify prodromal biomarkers to aid early diagnosis, to monitor disease progression, and to find a means for preventive treatment (Postuma and Berg, 2019; Videnovic *et al*., 2020).

Isolated rapid-eye-movement sleep behavior disorder (iRBD) is recognized as the most reliable prodromal biomarker of synucleinopathies such as Lewy body dementia (LBD) and Parkinson’s disease (Schenck *et al*., 2013; Iranzo *et al*., 2014; Postuma *et al*., 2019). In patients with RBD, the characteristic atonia of voluntary muscles in rapid-eye-movement (REM) sleep is impaired, leading to the acting out of dreams. Initially, patients start to vocalize, speak, and move complexly. As the disease progresses, patients may yell, fight, or jump out of bed, often injuring themselves or their sleeping partner (Schenck *et al*., 1986).

The mainstay of treatment in RBD is the prevention of injuries by modifying patients’ sleep environment and suppressing motor activity during sleep with muscle relaxants, usually clonazepam (Iranzo *et al*., 2016; Videnovic *et al*., 2020). Clonazepam immediately improves motor behavior during the first night of treatment. Being reluctant to initiate a long-term benzodiazepine treatment we introduced the use of melatonin as a therapy for patients with RBD (Kunz and Bes, 1997, 1999). Four common observations found in three different patient populations (Kunz and Bes, 1999, 2001; Kunz *et al*., 2004; Kunz and Mahlberg, 2010) pointed towards the circadian clock to be involved in the mode of action: 1. response occurred only when melatonin was administered within a narrow time span between 10 and 11 pm; 2. responders and non-responders were best differentiated by a stable clock-time of administration as opposed to flexible administration (e.g. “at bedtime” or “after meal”); 3. gradual response over weeks; 4. persisting effect after discontinuation of treatment. Based on these observations, we have developed a “chronobiotic protocol” for the use of melatonin treatment (Kunz, 2004). All our patients to be treated with melatonin are advised to follow this protocol since then. Although these effects may seem peculiar, melatonin is nowadays considered a first-line treatment in RBD, like clonazepam (Aurora *et al*., 2010; Videnovic *et al*., 2020).

Whereas reported rates of phenoconversion to clinical synucleinopathy rely on cohorts with a mixture of treatment strategies, primarily clonazepam (Fernandez-Arcos *et al*., 2016; Postuma *et al*., 2019; Galbiati *et al*., 2019), no large dataset has been reported for patients with iRBD treated with melatonin only. In this prospective cohort study, we aimed to determine the frequency of phenoconversion in two cohorts of patients with iRBD, one treated with the usual mixture of treatments and one with melatonin according to the chronobiotic protocol.

## Materials and methods

### Patients

Until August 2020 we diagnosed RBD in 282 consecutive patients according to the International Classification of Sleep Disorders (ICSD-3) using three night’s video-polysomnography (vPSG). Isolated RBD was present in 218 patients of whom 12 were diagnosed recently and by consequence, a follow-up examination was not yet performed (Fig. 1). Of the 206 patients evaluated, 167 patients were treated with melatonin according to our chronobiotic protocol, 39 were treated with the usual mixture of treatments.

**Figure 1.**
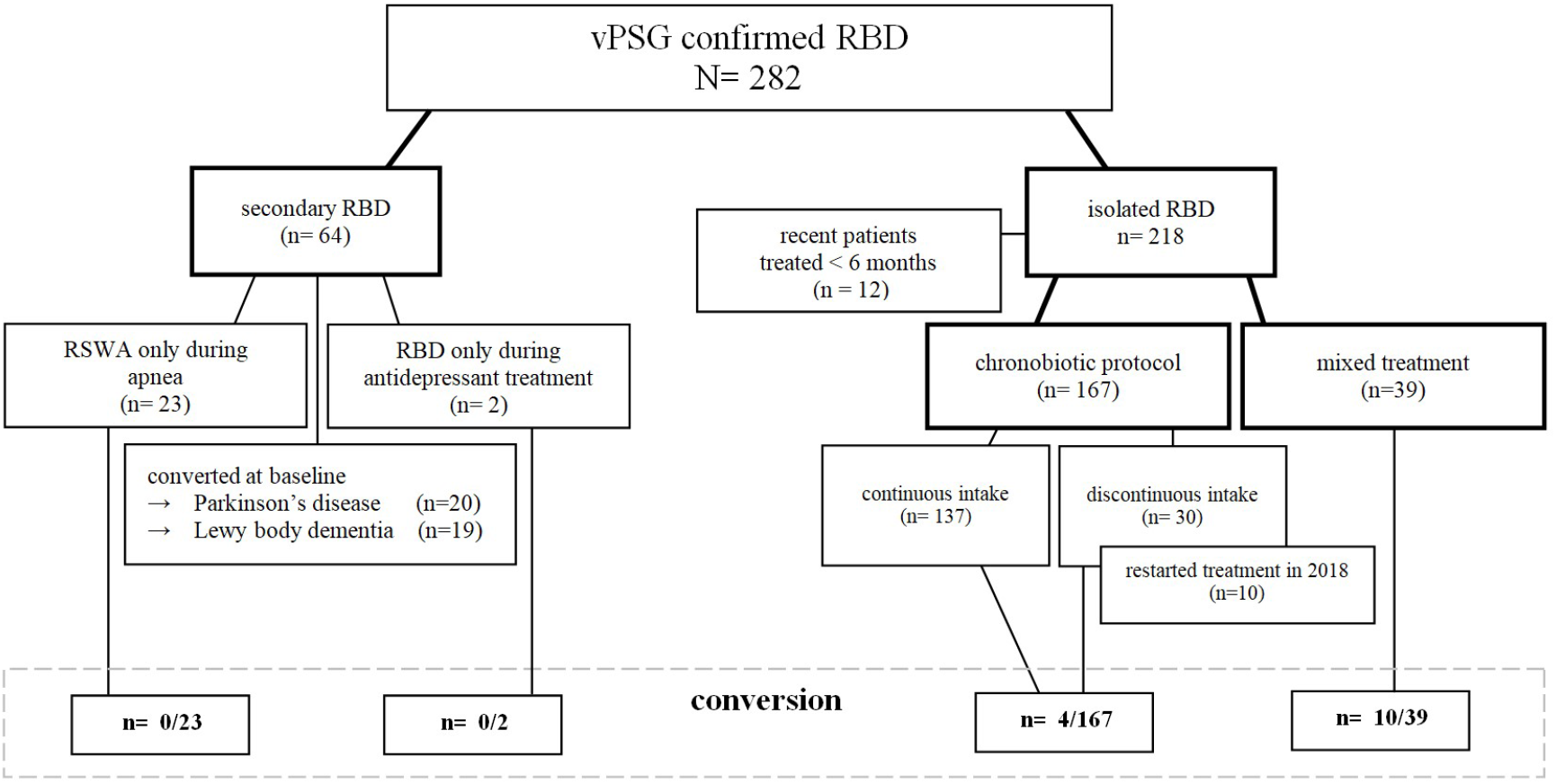
Patients with RBD Diagnosed 2008 – 2020. All RBD diagnoses confirmed by three night’s vPSG; “recent patients” with iRBD were diagnosed in 2020 and therefore without 6 months of melatonin treatment or follow-up including CERAD and UPDRS. Patients with “discontinued intake” stopped melatonin intake mostly after 6 months.

In our clinic, most patients are referred by a neurologist / general practitioner or contact us personally. They are diagnosed and redirected with a therapeutic recommendation. In patients with RBD we usually initiate treatment and patients are advised to administer melatonin on a long-term basis. In most of RBD patients our patient care stops, when symptoms are resolved. In those patients, who are reluctant towards medication, we used to recommend administration of melatonin for at least 6 months. Even though long-term melatonin treatment had been recommended to all of our patients, 39 iRBD patients had not been treated with melatonin for 6 or more months (“mixed treatment” group). This was because they discontinued melatonin therapy after concluding that “they were cured” because RBD symptoms did not reoccur after discontinuation of melatonin up to 3 months (n=13, “short-term melatonin” in Fig. 4). The remaining 26 patients (“various” in Fig. 4) experienced an initial worsening or treatment failure (melatonin stopped after 1-7 nights; n=16), and were hesitant to take melatonin or their treating physicians were reluctant to prescribe it (melatonin was stopped within 4 weeks; n=10). In those 26 patients the usual mixture of therapies was applied by their neurologist / general practitioner (e.g. clonazepam, other benzodiazepines, zolpidem, dopaminergic agents, mirtazapine, trazodone or no treatment).

The RBD-specific characteristics of our patients were compared in tabular form (Table 1) with three different kinds of representative samples (Fernandez-Arcos *et al*., 2016; Postuma *et al*., 2019; Galbiati *et al*., 2019).

**Table 1.** Generalizability of RBD Specifics.

|  | Postuma et al.,<br>2019 | Galbiati et al.,<br>2019 | Fernández-Arcos et<br>al., 2016 | Kunz et al.,<br>present study |  |
| --- | --- | --- | --- | --- | --- |
| Study design | multicenter | systematic review<br>and meta-analysis | single center | single center |  |
| Sample size N | 1280 | 3865 | 203 | 39 | 167 |
| Treatment | mixed | mixed | mixed | mixed | chronobiotic protocol |
| Male (%) | 83 | 78 (n=2444) | 80 | 72 | 78 |
| Age at estimated RBD onset (years) | – | – | md=63 (40–81) | 61 ± 12 (23–84) | 64 ± 9 (39–84) |
| Age at baseline/RBD diagnosis (years) | 66 ± 8 | 68 ± 3 (n=2444) | md=68 (50–85) | 66 ± 11 (30–86) | 69 ± 8 (41–86) |
| RBD severity: falling out of bed (%) | – | – | 77 | 64 | 62 |
| Obstructive sleep apnea (%) | 27 | – | 12 | 34 | 27 |
| Excessive daytime sleepiness (ESS) | – | – | 8.3 ± 4.7 | 7.4 ± 4.5 | 7.1 ± 4.0 |
| Beta blockers (%) | – | – | – | 26 | 33 |
| Antidepressants (%) | – | – | 26 | 31 | 18 |
| DaT-SPECT abnormal (%) | 44 (n=245) | – | – | 30 (n=10) | 35 (n=117<sup>c</sup>) |
| UPDRS-III | 6.26 ± 4.94 (n=57)<sup>a</sup><br>3.12 ± 3.82 (n=299)<sup>b</sup> | – | – | 1.0 ± 2.0 (n=17) | 2.1 ± 3.1 (n=81) |
| MMSE | 26.9 ± 3.4 (n=132)<sup>a</sup><br>28.1 ± 1.8 (n=375)<sup>b</sup> | – | – | 29.0 ± 0.9 (n=16) | 29.2 ± 1.0 (n=99) |
| Olfaction abnormal (%)<br>hyposmia<br>anosmia | 67 (n=628) | – | – | 47<br>47 (n=15) | 41<br>56 (n=96) |
| Constipation/gastrointestinal dysfunction (%) | 43 (n=830) | – | – | 43 (n=14) | 47 (n=38) |
| Urinary dysfunction (%) | 31 (n=687) | – | – | 57 (n=14) | 51 (n=37) |
| Sexual dysfunction (%) | 43 (n=276) | – | – | 63 (n=8) | 59 (n=29) |
| Average follow up (years) | 3.6 (max. 19) | 4.75 ± 2.43 | md=5 (0.1–17) | 3.1 ± 2.1 (0.3–7.6) | 4.2 ± 3.1 (0.6–21.7) |
| Conversion % (n) | 28 (352) | 31.9 (1235) | 34 (69) | 25.6 (10) | 2.4 (4) |
| Conversion-rate per year (%) | 6.3 | 6.7 | 6.8 | 8.3 | 0.6 |

Demographics and RBD-specific clinical data of our cohort and of three representative cohorts including neuromotor (UPDRS-III) and cognitive performance (Mini Mental Status Examination - MMSE), olfactory ability (Sniffin' Sticks), autonomic functioning (SCOPA-AUT), and dopamine-transporter (DaT)-SPECT. Numbers are given in percent, mean values with standard deviations, and medians (md) with ranges. Numbers are before melatonin treatment, except for DaTSCAN for which 40 patients were scanned after initiation of melatonin treatment. \*a: developed disease. \*b: disease free. \*c: 40/117 had DaT-SPECT during treatment.

### Chronobiotic Protocol

In our clinic we offer patients clinically suspected to have RBD to pass through a three night’s vPSG. Prior to the third night, we advise them to initiate a long-term melatonin treatment according to a chronobiotic protocol, which implies one major instruction: to administer melatonin “always-at-the-same-clock-time” once per day. This clock time should be established at 30 minutes prior to patient’s *habitual* bedtime. In cases where this fixed time cannot be kept, patients are instructed to skip melatonin intake that night. The rationale for this strict schedule is that since our initial pilot studies with RBD patients, we repeatedly observed that responders and non-responders were best distinguished by evaluating their sleep hygiene, i.e. stable vs. varying bedtimes and times of melatonin intake (summarized in Kunz, 2004). This clinical observation is in agreement with the fact that melatonin is known to feed back on the suprachiasmatic nucleus, the central pacemaker or masterclock (Gilette and McArthur, 1996; Gerdin *et al*., 2004). As a consequence, exogenous melatonin should be administered consistently within a rather narrow time span in order to gain optimal effects. Patients are informed that melatonin in RBD rarely exhibits effects during first days of treatment, rather effects occur within the first two weeks. Sometimes symptoms even worsen over the first days of administration, presumably because of transient initial sleep disruption due to immediate chronobiotic effects of melatonin, i.e. delaying or advancing the patient’s actual circadian phase (Lewy *et al*., 1992). In those patients, in whom melatonin does not show positive effects over the first three weeks of treatment, the time of administration is controlled referring to individual chronotype, e.g. in case of a mismatch with habitual bedtime.

### RBD Test Battery

When findings began to spread on the internet that iRBD represents a prodromal stage of clinical synucleinopathy, patients with RBD symptoms sought information on which stage of prodromal synucleinopathy they might be in. Since the end of 2015, we have therefore offered to all our new RBD patients a battery of examinations consisting of neurocognitive testing (CERAD-Plus, Schmid *et al*., 2014), neurologic/motoric evaluation of Parkinson’s severity (MDS-UPDRS-part III, Goetz *et al*., 2008), neurovegetative questioning (SCOPA-AUT, Visser *et al*., 2004), olfaction testing (Sniffin’ Sticks, Hummel *et al*., 1997), and dopamine transporter single-photon-emission computed tomography (DaT-SPECT, Djang *et al*., 2012)). CERAD-Plus, UPDRS-III, and SCOPA-AUT may be repeated annually. Over time, we expanded this initiative also to former patients being already under melatonin treatment (n=50), when they contacted us for follow-up.

Starting October 2018, we contacted all of our patients with iRBD for the evaluation of phenoconversion to clinical synucleinopathy. For treatment response, we specifically developed an RBD-symptoms severity rating scale called Ikelos-RS, after the god and personification of nightmares in the Greek mythology (inter-rater reliability: (correlation, Spearman’s rho) ρ=0.9, p<0.001; test-retest reliability: ρ=0.9, p<0.001; further details see Supplementary Methods).

All patients had provided written informed consent for their clinical data to be analyzed and published anonymously. The ethics committee of Charité - Universitätmedizin Berlin approved publication of the results of the post-hoc data analysis.

### Outcomes

The primary outcome was phenoconversion to clinical synucleinopathy after PSG-confirmed diagnosis, in a time-to-event analysis. Data for 206 evaluated patients were censored at last follow-up 2018 to 2020 (n = 183), or at the time when a patient was diagnosed as having phenoconverted (n = 14), had died (n = 8), or was lost to follow-up (n = 1). Phenoconversion was diagnosed at our site by means of a clinical neurological evaluation (criteria of Movement Disorders Society), supported by CERAD-Plus and UPDRS-III (performed in 141, respectively 153 of 167 patients under long-term melatonin treatment). Additionally, whenever the referring neurologists initiated specific treatment for parkinsonism/dementia, this was considered to be due to phenoconversion. Eight of 167 treated patients had died, and 11 could not be reached in person. In these 19 cases, spouses and referring physicians were contacted and asked if the patients had shown signs of parkinsonism/dementia. In only one patient, follow-up during prospective evaluation of phenoconversion after October 2018 was not possible. The last available follow-up was in 2017, five years after diagnosis, because the family had moved to an unknown new place of residence.

Secondary outcomes were variations over time in initial and follow-up scores on cognition (CERAD-Plus), neuromotor performance (UPDRS-III) and RBD symptom severity (Ikelos-RS, Clinical Global Impression (CGI)). In total, 1489 Ikelos-RS and CGI, 396 CERAD-Plus and 406 UPDRS-III scores were examined. Statistical analyses included only data from patients who at baseline had not yet taken melatonin and who underwent at least one follow-up examination during further treatment.

To analyze changes in RBD symptom severity over the first 6 months of treatment in more detail (Fig. 4), patients treated according to the chronobiotic protocol were classified into four subgroups according to known confounding factors with melatonin intake: use of beta blockers, use of antidepressants, melatonin intake not always-at-the-same-clock-time, and absence of these confounding factors. For analysis after this initial period, patients were re-classified into 2 subgroups, one with continued melatonin intake, and one with patients who stopped or interrupted melatonin at or shortly after this time. Those 39 patients being treated with the usual mixture of therapies were classified in two groups: one with positive initial response (“short-term melatonin”; improvement ≥ 2 on Ikelos-RS and CGI-S; n = 13) during melatonin treatment over 1 to 3 months (feeling “cured”) and one without positive response (“various”; n = 26) during less than 4 weeks melatonin treatment (n = 16) or no melatonin treatment (n = 10).

### Statistical analysis

Phenoconversion to clinical synucleinopathy was investigated in groups with or without long-term melatonin treatment using Kaplan-Meier survival analysis and the log-rank test. Hazard ratios and 95% confidence intervals were calculated using a Cox proportional-hazards model. In patients treated with melatonin, variation in cognitive (CERAD-Plus) and motor (UPDRS-III) performance over time was tested using linear mixed models. Model 1 analyzed each subtest (using z-scores) as dependent variable, treating “time category” as a fixed factor, “patient ID” as a random factor, and “continuous vs. discontinuous melatonin intake” and “normal vs. borderline/pathological DaTSPECT” as covariates. Based on the hypothesis that the clinically most impaired patients benefit most from treatment, subtests with significant results were categorized into low, medium, and high performers at baseline. This group factor was analyzed in model 2. Variations over time in RBD symptom severity (Ikelos-RS), both within and between subgroups, were analyzed using a random intercept model. Bonferroni post-hoc test was used for multiple testing corrections in all executed mixed models. Significance was defined as p-values less than 0.05. Analyses and modeling were implemented using IBM SPSS Statistics (version 23.0). Details of these analyses are provided in the supplementary material.

## RESULTS

A comparison of the RBD specifics of our patients with those of three representative cohorts suggests that our results are generalizable (Table 1). A remarkable exception, however, is the rate of phenoconversion to clinical synucleinopathy in our patients on long-term melatonin treatment according to chronobiotic protocol, which was extremely low: only 4 out of 167 patients (2.4%) converted after a mean follow-up of 4.2±3.1 years, implying a conversion rate of 0.6%/year. This is in sharp contrast to the phenoconversion rate in a) our 39 patients on mixed therapies, of whom 10 (25.6%) converted after a mean follow-up of 3.1±2.1 years, yielding a conversion rate of 8.3%/year and b) the three representative patient cohorts with mixed therapies used for comparison, in which the conversation rate ranged from 6% to 7%/year (Tab. 1, Fig. 2). In our cohort, treatment with melatonin for at least 6 months was associated with a reduced hazard of conversion of 0.07 (95% CI, 0.02-0.22; p< 0.001; Supplementary Tab. 1).

**Figure 2.**
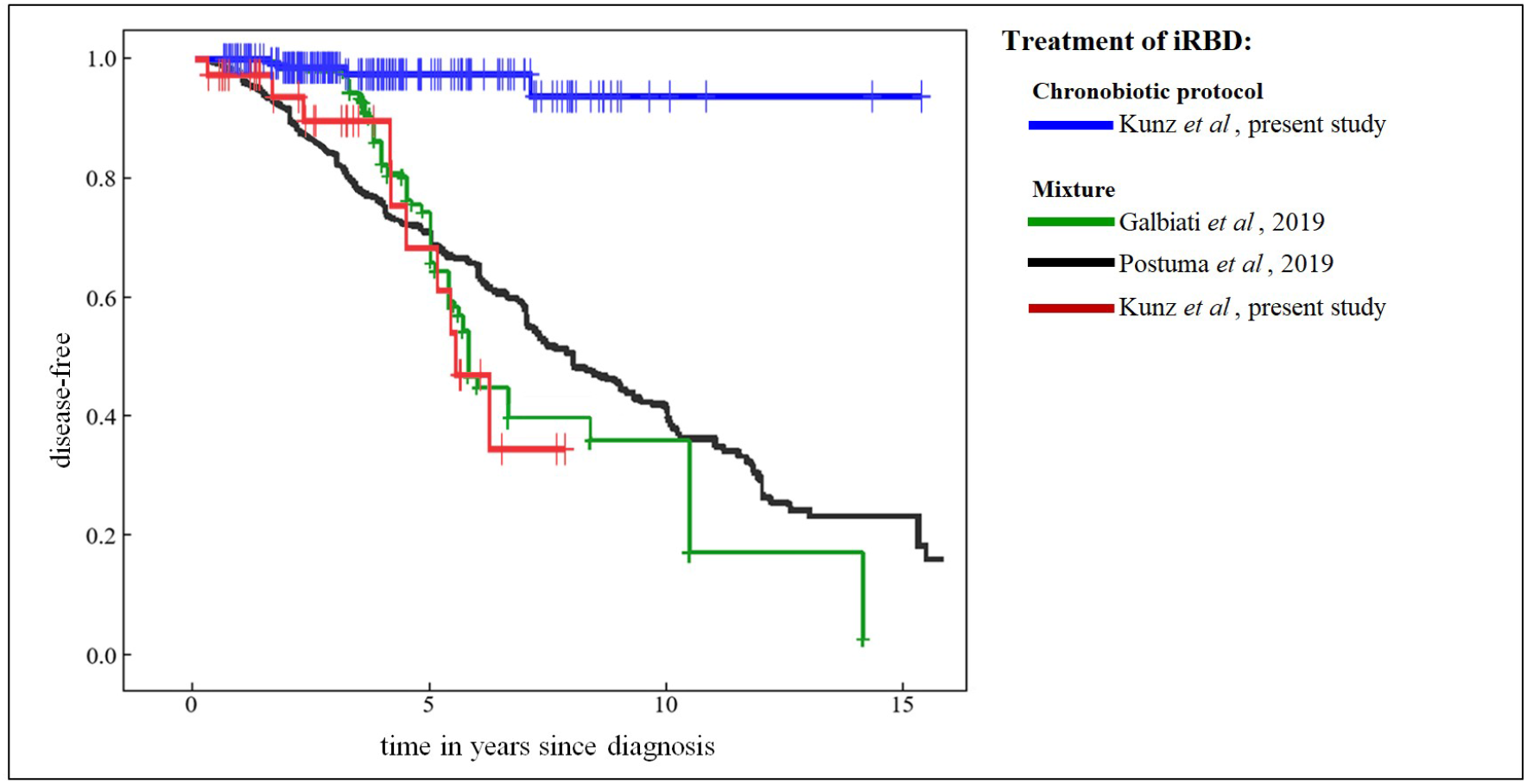
Phenoconversion from iRBD to Cinical Synucleinopathy. Kaplan-Meier curves, showing the cumulative disease-free survival. Right-censored events (vertical dashes) represent individual times of last follow-up, death or determination of conversion. To facilitate comparison, we depicted similar curves from two other published cohorts (Postuma *et al*., 2019; Galbiati *et al*., 2019 - with permission from Elsevier).

Moreover, cognitive performance – available from 91 patients before long-term melatonin treatment – did not decline over time (Fig. 3; Supplementary Tab. 2) and even improved on several subtests, with significant main effects seen for “Wordlist Encoding - verbal memory” (F_3,133_=7.33, p<0.001; Supplementary Tab. 3) and “Trail Making Test - Part A (TMT-A) – visual attention performance” (F_3,130_=5.02, p=0.003; Supplementary Tab. 4). In subtests for which significant results were observed, categorizing our cohort into low, medium, and high performers at baseline showed that low and medium performers improved over time in verbal memory (F_6,140_=4.94, p<0.001; Supplementary Tab. 5) and visual attention (F_6,135_=4.19, p=0.001; Supplementary Tab. 6), with low performers tending to achieve cognitive levels that were clinically inconspicuous (Fig. 3). Of the 16 patients with a mild neurocognitive disorder (NCD) at baseline, none had developed dementia or parkinsonism by the time of their last follow-up, two had worsened, three remained stable, and 11 had improved cognition. Of these 11 patients, eight did not fulfill the criteria for an NCD at last follow-up anymore. Of the remaining 151 patients without NCD at baseline, none developed an NCD during or after melatonin treatment.

**Figure 3.**
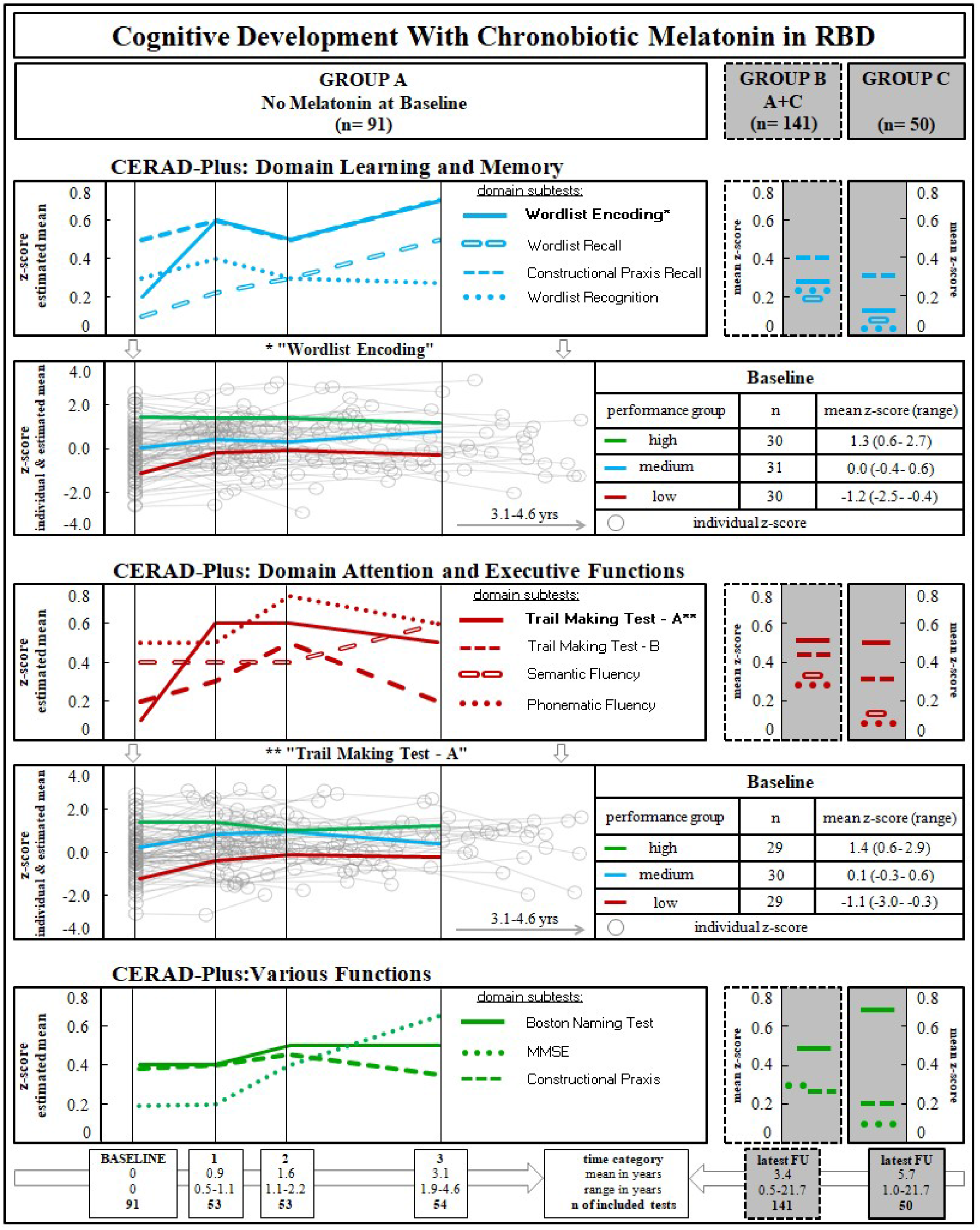
Cognitive Development with Chronobiotic Melatonin in RBD. CERAD-Plus subtests (total n = 396): results are z-scores of all 11 subtests; arranged in 2 cognitive domains and “various functions”; z-scores are based on comparison with standard population, taking into account gender, age and years of education; a z-score “0.0” represents average norm value. **Group A:** Baseline and all follow-up data of patients without melatonin intake at baseline examination are shown; follow-up times are categorized including roughly equal numbers of tests taken (at 0.9, 1.6, and 3.1 years after baseline). Individual examination results (grey circles, connected by linear interpolation lines) are presented for tests with statistically significant (“*” and “**”) time and group effects (“Wordlist encoding” and “Trail Making Test-A”);additional lines represent time courses of the categorized “high” (green), “medium” (blue), and “low” (red) performers at baseline. **Group B:** data at last follow-up of patients in groups A and C combined. **Group C:** data at last follow-up of patients who received melatonin already at baseline.

Neuromotor performance measured with UPDRS-III in 75 patients without melatonin at baseline (Supplementary Tab. 7) did not decline over time on long-term melatonin treatment either, but improved: the mean baseline score of 2.0 improved to 1.1 after 0.9 years and to 0.9 after 1.7 years, remaining stable at 0.7 after 3.5 years (F_3,104_=9.81, p<0.001; Supplementary Tab. 8). Again, the clinically most impaired performers improved more pronounced, with a mean total score of 6.6 at baseline and of 2.3 after 3.5 years (F_6,103_=13.33, p<0.001; Supplementary Tab. 9). The covariates “continuity of melatonin intake” and “DaT-SPECT classification” did not explain results in CERAD-Plus or UPDRS-III.

Fig. 4 shows that patients, who administered melatonin according to our chronobiotic protocol improved over first 4 to 6 weeks of treatment and remained stably improved. In contrast, patients administering melatonin, but not always at the same clock time, initially failed to show improvements in RBD symptom severity (F_5,1420_=10.25, p<0.001; Supplementary Tab. 10). Eleven of these patients reported an initial worsening of RBD symptoms during the first few days up to one week after starting melatonin treatment due to misinterpretation of the ambiguous prescription in the medication package leaflet to “administer melatonin after a meal, 1-2 hours before bedtime”. This resulted in intake that was too early, around 6 to 8 pm (i.e. after dinner). The remaining 14 patients in this category varied their time of intake by several hours, linking it to changing bedtimes. After instructions on proper timing were given again, RBD symptoms improved in all of these patients. Patients on concomitant beta blocker or antidepressant therapy responded more slowly and with less pronounced effects in the beginning. RBD symptoms leveled out at low Ikelos-RS values, with almost no patient reporting a complete disappearance of RBD symptoms. Improvement in Ikelos-RS was paralleled by a change in CGI severity (ρ=0.8, p<0.001). Of 167 patients on long-term melatonin treatment, 137 took melatonin at least 50% of the time between treatment initiation and last follow-up (max. 21.7 years), and 30 discontinued after 6 months. RBD symptoms did not worsen over time (F_1,812_=1.30, p=0.254; Supplementary Tab. 11) in any of these patients, including those who discontinued treatment after 6 months (Fig. 4, right panel).

**Figure 4.**
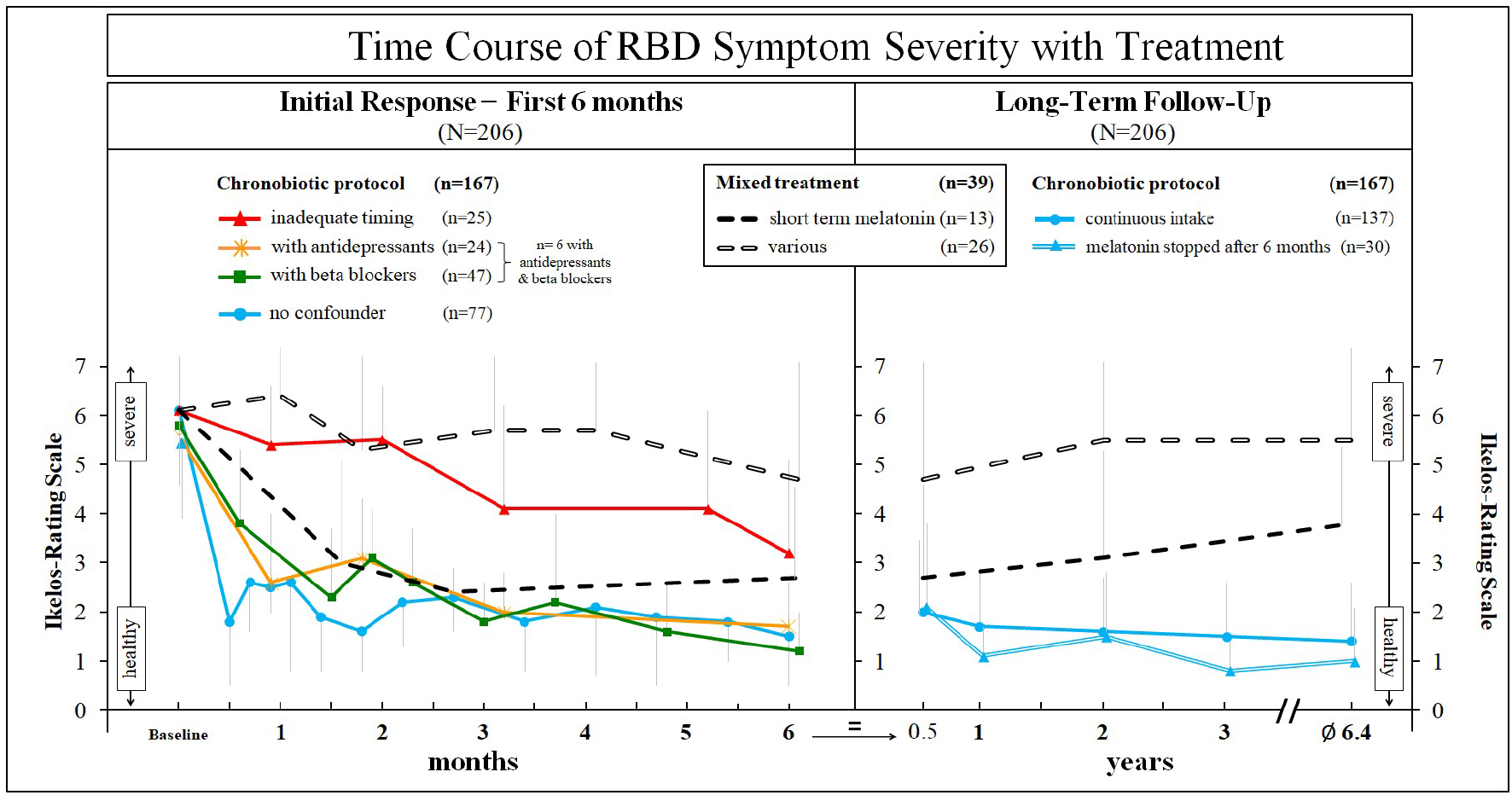
Time Course of RBD Symptom Severity with Treatment Chronobiotic Protocol: 2mg melatonin slow-release, ≥ 6 months, *always-at-the-same-clock-time*, between 10 and 11 pm - corrected for chronotype, melatonin intake to be skipped in case impossible during this time-span; **inadequate timing**: initial administration with varying clock-times (“before bedtime”) or too early (“after meal”); **Mixed Treatment - short term melatonin**: melatonin according to chronobiotic protocol but stopped after 1 to 3 months treatment; **various**: usual treatment of RBD at the advice of referring physician; **Left panel:** development of RBD symptom severity scores (Ikelos-RS) over the first 6 months; **Right panel:** symptom severity shown over subsequent years; for the chronobiotic protocol the former sub-groups are combined and now subdivided into a) one with continuous melatonin intake until last follow-up and b) one in which patients stopped melatonin intake after 6 months (patients felt “cured”). Ikelos-RS (total n = 1.498): observational periods were either the last 6 months (default), or since the last interview if less than 6 months; sum (range 0 to 7) of scores “frequency” of RBD symptoms (never = 0, ≤ 2 to 3 times a month = 1, 1 to 2 times a week = 2, 3 to 5 times a week = 3, daily = 4) and “severity” (single most severe acting out: speech or slight distal movements = 0, screaming or complex, non-aggressive movements = 1, complex movements with risk of injury = 2, leaving the bed = 3). Ikelos ratings based on bed partner interview only; 8 patients had no regular bed partner at all times; for these patients, reports of friends/relatives during periods of e.g. vacation or visits were included.

In the cohort of patients on mixed treatments acting out of dreams improved in 13 patients (Fig. 4; “short-term melatonin”) and administration of melatonin was stopped after 1 to 3 months because patients felt “cured” and considered a long-term medication with melatonin to be unnecessary. Symptoms partly returned in frequency and severity in follow-up and five patients converted (F_3,782_=3.57, p=0.014; Supplementary Tab. 12). Those patients without an initial improvement (Fig. 4, “various”) did not report changes in frequency or severity of RBD symptoms over time (e.g. a wearing-off of RBD symptoms).

## DISCUSSION

The main finding of our study is an exceptionally low rate of conversion to parkinsonism and dementia in the group of patients with iRBD who were under long-term melatonin treatment according to our chronobiotic protocol. With the seminal publication by Schenck *et al*. in 1996, initial evidence became available that iRBD was somehow linked to parkinsonism. Today it is recognized that iRBD represents the most reliable marker of prodromal state in the neurodegenerative process leading to clinical synucleinopathies, with a stable phenoconversion rate of 6% to 7% per year (Fernandez-Arcos *et al*., 2016; Postuma *et al*., 2019; Galbiati *et al*., 2019). Thus, the number of converters among 167 patients with iRBD over 4.2 years would be expected to have been 40 to 50, but in our cohort of patients we diagnosed only four, with either Parkinson’s disease or Lewy body dementia.

This result for our primary outcome is well supported by the results for our secondary outcome variables. Cognitive performance and motor functioning decline with healthy aging, and this process is accelerated with neurodegeneration (Terzaghi *et al*., 2013; Darweesh *et al*., 2017). Our patients treated with melatonin, however, not only did not experience a decline; they even improved in neuromotor and some subdomains of cognitive performance. Improvements were most pronounced in low performers (including patients diagnosed with NCD at baseline), who tended to reach performance levels that were near normal. Most interestingly, attention, which has been identified as a first-line cognitive indicator of developing LBD (Genier-Marchand *et al*., 2017), was significantly improved in low and medium performers. Assuming that the underlying neurodegenerative process has started at least at the onset of RBD and is already well underway, it is worth noting that in melatonin treated patients performance in nearly all cognitive subtests of the CERAD-Plus was on average still clearly above representative norm levels (i.e., z-score of zero) after many - here 8.7±5.3 - years after reported onset of RBD-symptoms.

All of our patients presented at baseline with involuntary falling out of bed or having attacked their bed partner (indicating a high severity of RBD symptoms) in the 3 months before RBD diagnosis. Within 2 weeks after starting melatonin treatment, however, while bed partners still noticed complex behavior, aggressive behavior had almost disappeared, which was objectivated by substantial improvement in Clinical Global Impression and RBD symptom severity. Improvements in RBD symptoms persisted for years even when melatonin was discontinued. This long-lasting improvement cannot be attributed to a wearing off of symptoms over time. Our group of patients being treated with the usual mixture showed no improvement over time. The time of treatment necessary for melatonin remains unclear and can only be estimated. Whereas patients who stopped melatonin administration after at least 6 months remained improved, patients who discontinued melatonin after 4 to 12 weeks worsened again with respect to RBD symptoms – though not to baseline levels - and converted at a similar rate to clinical synucleinopathy as those not treated according to a chronobiotic protocol.

Three aspects need to be considered when interpreting our findings: their generalizability, the mode of action of melatonin, and the limitations of our study.

### Generalizability

Our cohort represents the largest single center cohort on iRBD reported thus far and is comparable to other published cohorts with respect, first, to all major characteristics of patients with RBD (e.g. sex, age of onset of RBD symptoms, RBD severity, co-morbidities, co-medications) and, second, to olfactory deficit, abnormal motor symptoms, cognitive impairment, and abnormal DaT-SPECT, which have been identified as markers of an advanced state in the phenoconversion of patients with iRBD (Iranzo *et al*., 2017; Postuma *et al*., 2019). Third, in patients in our cohort who had not been treated with melatonin according to our chronobiotic protocol over the long term, the conversion rate was as one would expect from published data of patients with iRBD.

### Mode of Action

The present co-first line treatment for RBD, clonazepam, immediately improves RBD-related motor behavior, but the symptoms return within two nights after discontinuation (Fernandez-Arcos *et al*., 2016). In contrast, melatonin is not known to have any direct effects on motor behavior. Moreover, symptoms remain improved long after discontinuation of treatment, sometimes after years in those patients who stop administration of melatonin. Neuroprotective effects via melatonin (Reiter *et al*., 2013; Cardinali, 2019) and its receptors (Jockers *et al*., 2016) are to be studied.

The present results and our experience with melatonin indicate the known chronobiotic effects of melatonin which are highly time-dependent. Exogenous melatonin shifts the internal circadian phase according to a phase response curve and can be used in jetlag or to adjust late chronotypes to social time schedules (Lewy *et al*., 1992). Crucial in the context of RBD, however, might be a second, yet under-appreciated chronobiotic effect of melatonin: its darkness promoting property (Utiger, 1992; Dawson and Armstrong, 1996). When administered during its endogenous rise in the evening, melatonin does not shift phase, but strengthens the coordination of nightly circadian factors, with consequences such as promoting wake-related activities in nocturnal species and sleep-related activities in diurnal species. The sleep stage predominantly driven and modulated by the circadian timing system is that of REM sleep (Dijk *et al*., 1995; Bes *et al*.; 1996; Saper *et al*., 2005). The timing of impulses to this system seems decisive and, once established, needs to be kept “always-at-the-same-clock-time”. In contrast, administering melatonin at an inappropriate time induces phase shifts resulting in desynchrony of internal rhythms (Samel *et al*., 1991; Monk *et al*., 2000). Some of our patients reported initial worsening of RBD symptoms (e.g. jumping out of bed 3 nights in a row), most likely due to inadequate timing with respect to their individual circadian phase. Of the four patients who phenoconverted despite long-term melatonin treatment, one administered melatonin strictly at bedtime, which included around 10 days per month melatonin at 8 a.m. because of daytime sleep after nightshift work. Another patient who converted shifted time of administration once per week for 3 hours due to occupational constraints. Inadequate timing of melatonin seems likely not only to fail in improvement, but rather seems to worsen symptomatology due to desynchronization. Those patients who properly administered melatonin according to our chronobiotic protocol but only for short-term, experienced a reappearance of their RBD symptoms over time. The observation that this group of patients converted at a similar rate as patients with the other known symptomatic mixture of treatment seems to indicate a necessity of at least 6 months melatonin treatment.

Only recently, various reports suggested sleep and the circadian timing system to be causally involved in neurodegenerative mechanisms including synucleinopathy (Leng *et al*., 2020; Hablitz *et al*., 2020). Over the last decade the previously unknown glymphatic system was described, elegantly explaining clearance of metabolic waste including synuclein from the brain during sleep (Xie *et al*., 2013; Holth *et al*., 2019; Fultz *et al*., 2019). Only this year, data in mice show that glymphatic influx and clearance exhibit endogenous, circadian rhythms (Hablitz *et al*., 2020). Circadian abnormalities and flattening of circadian rhythms have repeatedly been proven in patients with Parkinson’s disease (Videnovic *et al*., 2014; Leng *et al*., 2020). Thus, the assumption that a synchronizing strengthening of circadian rhythms via light and melatonin – the hormone of darkness – may ameliorate neurodegenerative processes per se is tempting (Kunz, 2017; Videnovic *et al*., 2017).

Another interesting link exists between melatonin and Parkinson’s disease. Patients treated with the beta2-adrenoreceptor antagonist propranolol are at an increased risk to develop Parkinson’s disease, whereas patients treated with the beta2-adrenoreceptor agonist salbutamol are at a reduced risk (Mittal *et al*., 2017). As has been known for decades, lipophilic propranolol blocks melatonin secretion from the pineal gland via beta receptors (Cowen *et al*., 1985). Suppression of melatonin with beta blockers predominantly affects REM sleep, which can be reversed by exogenous melatonin (van den Heuvel *et al*., 1997). These mechanisms might explain the confounding effect of beta blockers in the first weeks of melatonin treatment in the observations presented here. The reported increased risk to develop Parkinson’s disease is small in numbers. This may indicate that betablockers do not increase risk to develop Parkinson’s disease per se, rather an ongoing process of neurodegeneration might be accelerated by reduced melatonin levels mediated by betablockers.

Two facts challenge our results: No previous research on the use of melatonin to treat RBD has reported similar effectiveness as observed in our cohort, and no evidence exists that the approximately 60 million Americans – predominantly the elderly – who have been using melatonin on a nightly basis at the crest of the “melatonin madness” in the mid-nineties (Reppert and Weaver, 1995) have experienced reduced rates of parkinsonism. Melatonin has been sold worldwide for the past 25 years as a hypnotic to be administered in connection with clock time independent events (e.g. “after a meal”, “at bedtime”). Many, if not most people who administered melatonin will therefore not have adhered to a schedule based strictly on clock time. Another factor of importance is dosage. Because melatonin influences its own receptor (Gerdin *et al*., 2004), it is important to have a melatonin-free period over the day. Supraphysiologic melatonin doses, especially in low metabolizers, prevent the absence of melatonin during the day and could induce insensitivity in melatonin receptors the next evening (Braam *et al*., 2010).

### Limitations

This is an observational study and the validity of observer-dependent results might therefore be questioned. Although all RBD symptom severity ratings were performed unblinded, they were based on bed-partner observation only and the changes in symptom severity were substantial. Symptoms such as frightening aggressiveness several times per week improved in almost all patients long-term treated with melatonin, replaced by milder symptoms like speaking aloud and slight movements occurring every other week. With regard to the primary outcome, phenoconversion also has been evaluated by referring neurologists, who were unaware of a possible disease-modifying effect of melatonin. Furthermore, low performers and patients suffering from NCD at baseline profited most according to objective neurocognitive tests. This makes systematic error in motor and cognitive performance scoring unlikely.

In conclusion, the present results strongly suggest that well-timed, long-term administration of melatonin to patients with iRBD is associated with reduced phenoconversion to parkinsonism and dementia and with improvement of specific clinical symptoms of prodromal synucleinopathies. Thus, melatonin may be associated with disease-modification in the prodromal state of synucleinopathies.

## Data Availability

Data are available from the corresponding author on request.

## Notes

### Competing Interest Statement

The authors have declared no competing interest.

### Clinical Trial

DRKS00023534

### Funding Statement

No funding.

### Author Declarations

Ethical committee of Charite - Universitaetsmedizin Berlin

