## Supplement for "Conversion to Parkinsonism and Dementia in REM-Sleep Behavior Disorder Using the Chronobiotic Melatonin"

#### Supplementary Material

#### Table of Contents | Page Number

1. Methods
- 1.1 Procedure history 2
- 1.2 Patients 2
- 1.3 Staging of synucleinopathy 3
  - 1.3.1 Neurocognitive testing 3
  - 1.3.2 Neurologic/motoric evaluation 3
  - 1.3.3 Neurovegetative questioning 3
  - 1.3.4 Olfaction testing 4
  - 1.3.5 Dopamine transporter density 4
  - 1.3.6 Phenoconversion 4
- 1.4 RBD symptom severity Rating Scale – Ikelos-RS 4
- 1.5 Polysomnography 4
- 1.6 Statistical methods 5

2. Results
- 2.1 Survival analysis 6
  - **Table 1: Results Cox's proportional-hazards model** 6
- 2.2 CERAD-Plus 6
  - 2.2.1 Statistical analysis 6
    - **Table 2: Descriptive statistics "CERAD-Plus"** 7
  - 2.2.2 Results model 1 7
    - **Table 3: Results mixed model 1 "CERAD-Plus:Wordlist encoding"** 7
    - **Table 4: Results mixed model 1 "CERAD-Plus:Trail Making Test – Part A"** 8
  - 2.2.3 Results model 2 9
    - **Table 5: Results mixed model 2 "CERAD-Plus:Wordlist encoding"** 9-10
    - **Table 6: Results mixed model 2 "CERAD-Plus:Trail Making Test – Part A"** 10-11
- 2.3 UPDRS-III 11
  - 2.3.1 Statistical analysis 11
    - **Table 7: Descriptive statistics "UPDRS-III"** 11
  - 2.3.2 Results model 1 12
    - **Table 8: Results mixed model 1 "UPDRS-III"** 12
  - 2.3.3 Results model 2 13
    - **Table 9: Results mixed model 2 "UPDRS-III"** 13-14
- 2.4 Ikelos-RS/CGI 14
  - 2.4.1 Validation 14
  - 2.4.2 Statistical analysis 14
  - 2.4.3 Results model A 14
    - **Table 10: Results mixed model A "Ikelos-RS"** 14-21
  - 2.4.4 Results model B 22
    - **Table 11: Results mixed model B "Ikelos-RS" (I)** 22-23
    - **Table 12: Results mixed model B "Ikelos-RS" (II)** 24-27

References for Supplement 28

### 1. Methods

#### 1.1 Procedure history

The Clinic for Sleep- & Chronomedicine at St. Hedwig-Hospital in Berlin, Germany, was founded in 2008, with diagnosis and treatment initiation in neurologic/psychiatric sleep disorders as major aim. Patients are referred or initiate contact themselves to our outpatient clinic. Diagnosis is performed in clinical interview (including a battery of psychometric testing), sometimes supported by several weeks of actigraphy. Depending on indication – referring to criteria of the International Classification of Sleep Disorders (ICSD-3; AASM, 2014) – patients may undergo diagnostic video-supported polysomnography (vPSG). After final diagnosis, treatments are initiated or proposed and patients return to their referring physician.

In 1995 we introduced the use of melatonin as a therapy in patients with REM sleep Behavior Disorder (RBD) (Kunz and Bes, 1997). Melatonin showed a gradual response over weeks, sometimes months, but also an outlasting effect after discontinuation of treatment, sometimes over years (Kunz and Bes, 1999; Kunz and Mahlberg, 2010). Since then, we have therefore recommended or initiated melatonin treatment in almost all of our patients with RBD. Those patients who questioned the need to continue long-term melatonin treatment after RBD symptoms had improved, were advised to discontinue after at least six months treatment and to contact again in case of reemergence of symptoms, or on demand.

Patients clinically suspected to have RBD are always advised by us to pass through a three night's vPSG. Prior to the third night, long-term melatonin treatment according to a chronobiotic protocol is initiated. This protocol basically implies one major instruction: to administer melatonin "always at the same clock time" once per day. This clock time should be established at 30 minutes prior to the *habitual* bedtime. In cases where this fixed time cannot be kept, patients are instructed to skip melatonin intake that night.

At the beginning of 2017 we started to reanalyze our records in order to evaluate systematically the course of treatment response in our patients, as well as confounding factors and response rates to melatonin. During this evaluation we observed that the rate of phenoconversion to clinical synucleinopathy in our patients treated with melatonin was much lower than what was usually reported in the literature for patients with similar characteristics. Thus, starting October 2018, we contacted all of our RBD patients for the evaluation of phenoconversion to clinical synucleinopathy. For treatment response we specifically developed a RBD-rating scale called Ikelos-RS, which we validated with Clinical Global Impression – CGI (Guy, 1976).

#### 1.2 Patients

Until August 2020 a total of 282 consecutive patients initially had been diagnosed with RBD. Thirty-nine patients had already converted to Lewy body dementia (LBD; n=19) or Morbus Parkinson (n=20) at the time of diagnostic PSG or first melatonin intake. None of our patients had developed Multiple System Atrophy (MSA). Twenty-five patients were diagnosed with secondary RBD: 23 were suffering from sleep apnea with RSWA and out-acting behavior occurring during breathing abnormalities only; 2 had developed RBD after antidepressant treatment, which resolved clinically completely after medication had stopped. Twelve patients – diagnosed since January 2020 – did not yet have a follow-up visit including neurocognitive testing and neurologic/motor evaluation after at least six months of melatonin treatment. None of these 37 RBD patients (25 secondary recontacted in 2020 and 12 new) had developed clinical synucleinopathy.

Finally, 206 patients were diagnosed with isolated RBD (iRBD) due to clinical interview and vPSG according to ICSD-3 criteria and included in our observational study. Five patients within this group were considered as with provisional RBD. They reported clear clinical RBD behavior, clinical evaluation showed absence of any other possible causal disorder/medication, and vPSG showed RSWA, but no complex behavior during REM sleep in video recordings.

Thirty-nine of 206 iRBD patients chose not to be treated with melatonin over at least six months. Reasons were: a) patients experienced an initial worsening or treatment failure (melatonin stopped after 1-7 nights; n=16); b) patients were hesitant to take melatonin or their treating physicians were reluctant to prescribe it (melatonin was stopped within 4 weeks; n=10); c) patients discontinued because melatonin showed good effects and discontinuation after up to 3 months did not result in reoccurrence of RBD-symptoms: patients "felt no need in continuing treatment" (n=13). In those 26 patients feeling not sufficiently treated with melatonin, the usual mixture of therapies was applied by their neurologist or general practitioner (e.g. clonazepam, other benzodiazepines, zolpidem, dopaminergic agents, mirtazapine, trazodone).

Of these 39 patients, 35 could directly be re-contacted in 2019/2020 for follow-up. Three patients had died but referring physician reported premortal physical/mental status, one patient could not be reached in person and other sources of information were not available. Consequently, the last available follow-up was included in our observation. 10/39 patients received beta blockers (Metoprolol (n=5) and Bisoprolol (n=5)) at baseline. 12 patients took one or more of the following antidepressant medication: Duloxetin (n=3), Venlafaxin (n=2), Sertralin (n=2), Agomelatine (n=1), Doxepin (n=1), Citalopram (n=1), Escitalopram (n=1), Mirtazapin (n=1), Opipramol (n=1), Paroxetin (n=1) and Trazodon (n=1).

The remaining 167 iRBD patients were long-term treated with melatonin, i.e. for at least six months, according to a chronobiotic protocol: they received a slow-release form 2mg/day, always at the same, individually determined clock time. Twenty-one patients temporarily increased melatonin doses to 4 and 8mg without any effect and returned to 2mg. In the light of a possible neuroprotective effect by melatonin in RBD, 10 of 30 patients who discontinued melatonin treatment after six months restarted melatonin treatment after re-contact in 2018. In none of these 30 patients RBD symptoms had worsened during melatonin-free period. Of the 167 patients, 147 could directly be recontacted in 2020 for regular follow-up. 11 could not be reached in person. In these cases, spouses and referring physicians were contacted and asked if the patients had recently shown signs of parkinsonism or dementia. Eight patients had died and the referring physician could report premortal physical / mental status. In 1 patient information was unavailable (family had moved to an unknown place of residence in 2017; the last available follow-up, five years after diagnosis/treatment initiation, was therefore used in our observation). 55/167 patients received beta blockers (Metoprolol (n=25), Bisoprolol (n=23), Nebivolol (n=5), Solatol (n=1), unknown (n=1)) at baseline. 30 patients took one ore more of the following antidepressant medication: Citalopram (n=7), Escitalopram (n=5), Venlafaxin (n=5), Duloxetin (n=3), Mirtazapin (n=3), Trimipramin (n=3), Amitriptylin (n=2), Clomipramin (n=2), Paroxetin (n=2), Agomelatine (n=1), Bupropion (n=1), Doxepin (n=1), Nortrilen (n=1) and Opipramol (n=1).

#### 1.3 Staging of synucleinopathy

Starting by the end of 2015, all of our iRBD patients were offered a staging of prodromal state in the synucleinopathy process. The test-battery consisted of neurocognitive testing, neurologic/ motoric evaluation of Parkinson severity, neurovegetative questioning, olfaction testing, as well as dopamine transporter single-photon emission computed tomography (DaT-SPECT). CERAD-Plus and UPDRS-III were repeated annually since.

##### 1.3.1 Neurocognitive testing

The extended German version of the Consortium to Establish a Registry for Alzheimer's Disease (CERAD-Plus, Schmid *et al.*, 2014) consisting of 11 different subtests, was used to measure cognitive performance on the domains language, memory, attention/praxis and orientation. Data entered by the patient are converted into z-values, based on a comparison with the standard population (CERAD: N=1100; age 49-92 years; Plus: N=604; age 55-88 years, Schmid *et al.*, 2014), taking into account gender, age and years of education. The CERAD-Plus was performed in 141 at least once after six months of melatonin treatment, with an average follow up of 3.4±2.9 years. Ninety-one patients were without melatonin treatment at initial examination. Fifty patients had started melatonin treatment prior to the initial CERAD-Plus-examination, implying that for them no CERAD-Plus was available at baseline.

##### 1.3.2 Neurologic/motoric evaluation

Presence of motor signs was assessed using the Movement Disorders Society Unified Parkinson Disease Rating Scale (MDS-UPDRS) part III (Goetz *et al.*, 2008). In the present study, 153 patients were tested within the UPDRS-III at least once after at least six months of melatonin treatment with an average follow up of 3.5±2.9 years. Seventy-five patients were without melatonin treatment at initial examination. Seventy-eight patients had started melatonin treatment before the initial examination, implying that for them no UPDRS-III was available at baseline.

##### 1.3.3 Neurovegetative questioning

Autonomic dysfunction was evaluated since 2018 in 38 patients without melatonin at baseline examination using the Scales for Outcomes in Parkinson's disease-Autonomic (SCOPA-AUT, Visser *et al.*, 2004), assessing gastrointestinal, urinary, cardiovascular, thermoregulatory, pupillomotor and sexual symptoms.

##### 1.3.4 Olfaction testing

Olfactory abilities were measured once in 130 patients via Sniffin‘ Sticks Extended Test (Hummel *et al.*, 1997), of which 96 patients did not receive melatonin at baseline. The results of three subtests (threshold, discrimination and identification test) were added up to a total score, which was classified as ≤15.5: anosmia, <31: hyposmia and ≥31: normosmia.

##### 1.3.5 Dopamine transporter density

Patients underwent DaT-SPECT after intravenous injection of 130 MBq $^{123}$I-FP-CIT, in order to evaluate reduced presynaptic dopamine transporter binding. Striatal-to-occipital cortex uptake ratios and z-scores (judged to be reduced when more than 2 SD) were calculated for caudate nucleus and anterior and posterior putamen. The interpretation of FP-CIT SPECT was based on visual assessment of the reconstructed SPECT images by a nuclear medicine expert. Visual reading was supported by semiquantitative BRASS-analysis. Based on both statistical and visual analysis scans were evaluated as “inconspicuous” (clear, left-right symmetric delineation of both striata), “pathologic” (distinct uniform or asymmetric reduction) or “borderline” with respect to the presence of Parkinson's disease (Zubal *et al.*, 2007; Djang *et al.*, 2012). In total, 117 out of 167 patients underwent DaT-SPECT, from which 77 without melatonin at baseline conditions.

##### 1.3.6 Phenoconversion

In the cohort of patients being long-term treated with melatonin, phenoconversion to Morbus Parkinson, LBD or MSA was diagnosed on the basis of clinical neurological evaluation performed by our site. Decision was supported by CERAD-Plus and UPDRS-III. In those patients unable to be reevaluated at our site – due to death prior to recontact in 2018/2019/2020 (n=8), or not being interested in specific evaluation when contacted in 2019/2020 (n=11) – diagnosis of phenoconversion was decided on the basis of the referring neurologist´s evaluation. Almost all of the patients (91%) were under supervision of a referring neurologist. In remaining 1 patient we were unable to identify current address or physician. There, last follow-up available was included. Neurologists were not advised as to early or late start of specific antiparkinsonian / antidementia treatment. Whenever neurologists initiated such treatment this was considered as due to phenoconversion.

Neurocognitive Disorder (NCD) was defined according to the DSM-5 criteria (APA, 2015), i.a. subjective complaint about cognitive abilities and objective cognitive impairments (at least 2 subtests in CERAD-Plus with a z-score below -1.3; Aebi *et al.*, 2002). Out of 141 patients who were tested within the CERAD-Plus at least once after 6 months of melatonin treatment, 16 patients fulfilled the criteria for NCD at baseline examination.

#### 1.4 RBD symptom severity Rating Scale – Ikelos-RS

RBD symptom severity was rated by a clinician based on interview with patient’s bed partner only (mostly personal or by phone-call; when symptom severity was unchanged, patients report of bedpartner`s observation was accepted). Observational periods were either the last 6 months (default), or since the last interview if less than 6 months. In case a bed-partner did not exist (n=8), short-term observations by friends or families during visits or joined vacations were taken with shorter observational periods. The Ikelos-RS was filled out at baseline (approximately at the time of vPSG) and at every follow-up. Severity of RBD symptoms was evaluated on two dimensions: On the first scale “frequency” (running from 0 to 4), the occurrence of symptoms is estimated from never (=0), to ≤2-3 times a month (=1), 1-2 times a week (=2), 3-5 times a week (=3) and daily (=4). The second scale “expression” (running from 0 to 3), describes the form of the most severe manifestation of RBD symptoms from speech or slight distal movements (=0) to screaming or complex, non-aggressive movements (=1), complex movements with risk of injury (=2) and leaving the bed (=3). Both ratings are added to a total score (range 0-7) and are supplemented by two components of the Clinical Global Impression (CGI-Severity Scale, CGI-S, and CGI-Improvement Scale, CGI-I; Guy, 1976). Validation was performed with regard to convergent validity, interrater- and test-retest reliability (see lower section 2.4.1).

#### 1.5 Polysomnography

All patients underwent full night video-supported polysomnography (vPSG) on three consecutive nights. The first night was intended for adaptation to the laboratory conditions and the following two nights for diagnostic purposes. In case of RBD diagnosis, long-term treatment with melatonin was started on the third night.

PSG was performed with digital recording systems (predominantly Monet 24-CPU, TMS International, Enschede, The Netherlands or Sagura Polysomnograph 2000, Mühlheim, Germany, with Rembrandt version 7.5 software, Medcare Automation, Amsterdam, The Netherlands). It included electro-encephalographic (EEG) monitoring (sample rate 160 Hz, high-pass filter 0.3 Hz, low-pass 70 Hz) with the following 10-20-system based derivations: frontal region (F3, F4), central region (C3, C4) and occipital region (O1, O2), each referred to the contra-lateral mastoid (A1, A2). In addition, further continuous measurements were performed: Electro-oculography (sample rate 200 Hz, filter settings 0.3-70 Hz), bipolar surface EMG of m. mentalis and mm. tibiales anterior (sample rate 200 Hz, filter settings 10-120 Hz), electrocardiography (ECG), deep body temperature (rectal thermistor), respiratory flow (nasal-oral thermistor or nasal pressure sensor), respiratory effort of chest and abdomen (induction-plethysmography) and oxyhemoglobine saturation measured at the index finger. Snoring sounds were detected by either a microphone attached to the skin of the throat or derived from a bandpass (30-70 Hz) filtered air flow pressure signal. A video (infra-red, 25 fps resolution) and audio recording (PCM, 8 kHz, 8 bit, mono) was performed synchronously with all PSG measurements. Bio-calibration was performed prior to the start of each recording to ensure adequate signal quality. Each recording took place in a dark, low-noise single-bed room.

##### 1.6 Statistical methods

Scores of cognitive and motor examinations in patients from baseline to follow up tests were investigated with linear mixed model analyses. Only measurements of patients with at least one follow up after baseline were included and allocated to three different, consecutive time categories with comparable group sizes.

In model 1 it was tested whether changes in cognitive or motor scores could be detected over time. Within model 2 it was investigated whether performance in follow up examinations was dependent on the baseline level of cognitive or motor performance, testing the hypothesis that treatment with melatonin results in a normalization of eventual impairments, i.e. with clinically most impaired patients profiting most from treatment.

The validation of the Ikelos-RS within the present publication included evaluation of convergent validity within a bivariate correlation analysis (Spearman's rho) with Ikelos-RS and CGI-S. Spearman's rank correlation coefficient was also used to measure test-retest and interrater reliability. Further validation using i.a. videometric data and RSWA are part of the PhD thesis of one of the authors (SS) and will be reported elsewhere.

The Ikelos-RS total score is, strictly speaking, an ordinal measure. However, to allow robust statistical modelling and because higher Ikelos-RS-values are associated with more pronounced symptoms, a linear mixed model was preferred over a cumulative link mixed model for ordinal data to test time and group effects. In previous studies, it was shown that analyzing ordinally scaled data with parametric methods did not result in high type I error rates (Norman, 2010). Therefore, changes in RBD symptom severity and differences over time between different groups, categorized regarding therapy, confounding co-medication, inadequate timing or continuity of melatonin intake, were investigated within a random intercept model.

Bonferroni post-hoc test was used for multiple testing correction in all executed mixed models.

Within each model, p values less than 0.05 were evaluated to be significant. All statistical analyses were performed in IBM SPSS Statistics for Windows, version 23.0 (IBM Corp., Armonk, NY).

#### 2. Results

##### 2.1 Survival analysis

Log-rank test indicated a significant difference between the survival curves of patients with chronobiotic protocol (n=167) vs. mixed treatment (n=39) with melatonin ($\chi^2$=35.43, p<0.001).

Cox's proportional-hazards model was applied using group classification (chronobiotic protocol vs. mixed treatment) as an explanatory variable. Results indicate that there is strong evidence that group classification is associated with the length of survival, showing that being treated with melatonin reduces the hazard by a factor of 0.07.

**Table 1: Results Cox's proportional-hazards model**

| | | | | | | | 95 % CI for Exp(B) | |
|---|---|---|---|---|---|---|---|---|
| | B | SE | Wald | df | Sig. | Exp(B) | Lower Bound | Upper Bound |
| GROUP | -2.696 | .597 | 20.377 | 1 | .000 | .067 | .021 | .217 |

##### 2.2 CERAD-Plus

###### 2.2.1 Statistical analysis

###### 2.2.1.1 Implementation

The CERAD-Plus was performed at least once after six months of melatonin treatment with an average follow up of 3.4 ± 2.9 years (range: 0.5-21.7yrs) in 141/167 iRBD patients. For further analyses, only those who were untreated at baseline and examined cognitively multiple times (n=91) were included, with 160 follow-up examinations compared at three different time points (=TIMECATEGORY) to cognitive performances at baseline.

###### 2.2.1.2 Model specification

In model 1, analyses were executed for each subtest of the CERAD-Plus (using z-scores) as a dependent variable, treating TIMECATEGORY as a fixed, patient ID (=ID) as a random factor and continuity of melatonin intake (=MEL; "continuously" vs. "discontinuously") as well as DaTSPECT results (=DaT; "normal" vs. "borderline" vs. "pathological" scan) as covariates.

Within model 2, a group factor (=COGNITIONGROUP) was added as a fixed factor for the statistically significant tests from model 1 ("Wordlist encoding" and "Trail Making Test – part A"). Depending on the cognitive performance shown at baseline, patients were categorized into a group of "low", "medium" or "high" performer for each test separately. Additionally, the interaction term of TIMECATEGORY *COGNITIONGROUP was included as a fixed factor besides the variables defined in model 1.

###### 2.2.1.3 Descriptive statistics

Means and standard deviations with ranges in years of the follow up duration for the total and sub-cohort as well as the definition of TIMECATEGORIES, the number of re-tests and mean z-scores with ranges of the different COGNITIONGROUPS are characterized as follows:

**Table 2: Descriptive statistics “CERAD-Plus”**

| Follow-up durations and re-examinations | | | Linear mixed model | | | | | |
|---|---|---|---|---|---|---|---|---|
| Follow up of total cohort (yrs) | Follow up of included sub-cohort (yrs) | Number of re-tests | TIME C. 1 (yrs) | TIME C. 2 (yrs) | TIME C. 3 (yrs) | COGNITIONGROUP | | |
| | | | | | | low | medium | high |
| 3.4 ± 2.9<br>(0.5-21.7)<br>n=141/167 | 2.1 ± 1.2<br>(0.5-4.6)<br>n= 91/167 | 1x = 45<br>2x = 25<br>3x = 19<br>4x = 2<br>n= 160 | 0.9<br>(0.5-1.1)<br>n= 53 | 1.6<br>(1.1-2.2)<br>n= 53 | 3.1<br>(1.9-4.6)<br>n= 54 | Substest: “Wordlist encoding” | | |
| | | | | | | -1.2<br>(-2.5- -0.4)<br>n=30 | 0.0<br>(-0.4–0.6)<br>n=31 | 1.3<br>(0.6-2.7)<br>n=30 |
| | | | | | | Subtest: “TMT-A” | | |
| | | | | | | -1.1<br>(-3.0- -0.3)<br>n=29 | 0.1<br>(-0.3-0.6)<br>n=30 | 1.4<br>(0.6-2.9)<br>n=29 |

##### 2.2.2 Results model 1 (CERAD-Plus)

**Table 3: Results mixed model 1 “CERAD-Plus: Wordlist encoding”**

| Tests of Fixed Effects | | | | |
|---|---|---|---|---|
| Source | Num. df | Den. df | F | Sig. |
| Intercept | 1 | 69.682 | .003 | .956 |
| TIMECATEGORY | 3 | 133.748 | 7.330 | .000 |
| MEL | 1 | 75.339 | 2.098 | .152 |
| DaT | 1 | 69.673 | 3.751 | .057 |

| Estimates | | | | | |
|---|---|---|---|---|---|
| | | | | 95% CI | |
| TIME-CATEGORY | Mean | Std. Error | df | Lower Bound | Upper Bound |
| Baseline | .167 | .125 | 102.988 | -.081 | .415 |
| 1 | .564 | .141 | 140.949 | .285 | .843 |
| 2 | .528 | .146 | 150.549 | .240 | .817 |
| 3 | .677 | .153 | 160.049 | .375 | .978 |

| Pairwise Comparisons | | | | | | | |
|---|---|---|---|---|---|---|---|
| | | | | | | 95% CI | |
| (I) TIME-CATEGORY | (J) TIME-CATEGORY | Mean Difference (I-J) | Std. Error | df | Sig. | Lower Bound | Upper Bound |
| Baseline | 1 | -.397* | .120 | 129.018 | .007 | -.717 | -.077 |
| | 2 | -.361* | .125 | 131.437 | .027 | -.697 | -.026 |
| | 3 | -.510* | .133 | 135.047 | .001 | -.866 | -.154 |
| 1 | Baseline | .397* | .120 | 129.018 | .007 | .077 | .717 |
| | 2 | .036 | .147 | 137.968 | 1.000 | -.359 | .430 |
| | 3 | -.113 | .151 | 139.087 | 1.000 | -.517 | .292 |
| 2 | Baseline | .361* | .125 | 131.437 | .027 | .026 | .697 |
| | 1 | -.036 | .147 | 137.968 | 1.000 | -.430 | .359 |
| | 3 | -.148 | .153 | 137.141 | 1.000 | -.559 | .263 |
| 3 | Baseline | .510* | .133 | 135.047 | .001 | .154 | .866 |
| | 1 | .113 | .151 | 139.087 | 1.000 | -.292 | .517 |
| | 2 | .148 | .153 | 137.141 | 1.000 | -.263 | .559 |

##### Table 4: Results mixed model 1 "CERAD-Plus: Trail Making Test – Part A"

| Tests of Fixed Effects | | | | |
|---|---|---|---|---|
| Source | Num. df | Den. df | F | Sig. |
| Intercept | 1 | 67.279 | 4.929 | .030 |
| TIMECATEGORY | 3 | 130.461 | 5.024 | .003 |
| MEL | 1 | 73.664 | .966 | .329 |
| DaT | 1 | 67.193 | .665 | .418 |

| Estimates | | | | | |
|---|---|---|---|---|---|
| | | | | 95% CI | |
| TIME-CATEGORY | Mean | Std. Error | df | Lower Bound | Upper Bound |
| Baseline | .142 | .146 | 104.948 | -.148 | .432 |
| 1 | .591 | .169 | 147.888 | .257 | .925 |
| 2 | .619 | .172 | 151.083 | .280 | .959 |
| 3 | .492 | .181 | 160.936 | .135 | .850 |

| Pairwise Comparisons | | | | | | | |
|---|---|---|---|---|---|---|---|
| | | | | | | 95% CI | |
| (I) TIME-CATEGORY | (J) TIME-CATEGORY | Mean Difference (I-J) | Std. Error | df | Sig. | Lower Bound | Upper Bound |
| Baseline | 1 | -.449* | .151 | 125.586 | .021 | -.853 | -.044 |
| | 2 | -.477* | .154 | 127.476 | .014 | -.889 | -.065 |
| | 3 | -.350 | .164 | 131.750 | .209 | -.790 | .090 |
| 1 | Baseline | .449* | .151 | 125.586 | .021 | .044 | .853 |
| | 2 | -.029 | .183 | 134.845 | 1.000 | -.518 | .460 |
| | 3 | .099 | .189 | 136.900 | 1.000 | -.408 | .605 |
| 2 | Baseline | .477* | .154 | 127.476 | .014 | .065 | .889 |
| | 1 | .029 | .183 | 134.845 | 1.000 | -.460 | .518 |
| | 3 | .127 | .188 | 134.457 | 1.000 | -.377 | .632 |
| 3 | Baseline | .350 | .164 | 131.750 | .209 | -.090 | .790 |
| | 1 | -.099 | .189 | 136.900 | 1.000 | -.605 | .408 |
| | 2 | -.127 | .188 | 134.457 | 1.000 | -.632 | .377 |

##### 2.2.3 Results model 2 (CERAD-Plus)

**Table 5: Results mixed model 2 "CERAD-Plus: Wordlist encoding"**

| Tests of Fixed Effects | | | | |
|---|---|---|---|---|
| Source | Num. df | Den. Df | F | Sig. |
| Intercept | 1 | 68.260 | .371 | .544 |
| TIMECATEGORY | 3 | 140.448 | 9.558 | .000 |
| MEL | 1 | 83.506 | .039 | .844 |
| DaT | 1 | 68.629 | 5.578 | .021 |
| COGNITIONGROUP | 2 | 68.743 | 48.417 | .000 |
| TIMECATEGORY*COGNITIONGROUP | 6 | 140.042 | 4.936 | .000 |

| Estimates (COGNITIONGROUP*TIMECATEGORY) | | | | | | |
|---|---|---|---|---|---|---|
| | | | | | 95 % CI | |
| COGNITIONGROUP | TIMECATEGORY | Mean | Std. Error | df | Lower Bound | Upper Bound |
| low | Baseline | -1.145 | .162 | 138.223 | -1.465 | -.825 |
| | 1 | -.156 | .207 | 177.647 | -.565 | .253 |
| | 2 | -.060 | .201 | 173.918 | -.458 | .337 |
| | 3 | -.341 | .212 | 170.249 | -.760 | .078 |
| medium | Baseline | .046 | .139 | 138.417 | -.229 | .321 |
| | 1 | .385 | .153 | 159.896 | .083 | .687 |
| | 2 | .252 | .178 | 178.399 | -.099 | .602 |
| | 3 | .814 | .169 | 165.084 | .481 | 1.147 |
| high | Baseline | 1.372 | .146 | 136.912 | 1.083 | 1.662 |
| | 1 | 1.369 | .189 | 178.513 | .996 | 1.742 |
| | 2 | 1.418 | .187 | 174.071 | 1.049 | 1.788 |
| | 3 | 1.221 | .227 | 184.677 | .774 | 1.668 |

| Pairwise Comparisons (COGNITIONGROUP*TIMECATEGORY) | | | | | | | | |
|---|---|---|---|---|---|---|---|---|
| | | | | | | | 95% CI | |
| COGNITIONGROUP | (I) TIME-CATEGORY | (J)TIME-CATEGORY | Mean Difference (I-J) | Std. Error | df | Sig. | Lower Bound | Upper Bound |
| low | Baseline | 1 | -.989* | .217 | 133.703 | .000 | -1.572 | -.407 |
| | | 2 | -1.085* | .212 | 133.835 | .000 | -1.654 | -.516 |
| | | 3 | -.804* | .223 | 147.917 | .003 | -1.402 | -.207 |
| | 1 | Baseline | .989* | .217 | 133.703 | .000 | .407 | 1.572 |
| | | 2 | -.095 | .257 | 152.540 | 1.000 | -.781 | .591 |
| | | 3 | .185 | .271 | 164.326 | 1.000 | -.538 | .909 |
| | 2 | Baseline | 1.085* | .212 | 133.835 | .000 | .516 | 1.654 |
| | | 1 | .095 | .257 | 152.540 | 1.000 | -.591 | .781 |
| | | 3 | .281 | .248 | 145.077 | 1.000 | -.384 | .945 |
| | 3 | Baseline | .804* | .223 | 147.917 | .003 | .207 | 1.402 |
| | | 1 | -.185 | .271 | 164.326 | 1.000 | -.909 | .538 |
| | | 2 | -.281 | .248 | 145.077 | 1.000 | -.945 | .384 |
| medium | Baseline | 1 | -.339 | .164 | 123.783 | .243 | -.779 | .100 |
| | | 2 | -.206 | .187 | 132.283 | 1.000 | -.707 | .295 |
| | | 3 | -.768* | .179 | 139.158 | .000 | -1.247 | -.289 |
| | 1 | Baseline | .339 | .164 | 123.783 | .243 | -.100 | .779 |
| | | 2 | .133 | .202 | 141.299 | 1.000 | -.407 | .674 |
| | | 3 | -.429* | .191 | 141.718 | .157 | -.939 | .082 |
| | 2 | Baseline | .206 | .187 | 132.283 | 1.000 | -.295 | .707 |
| | | 1 | -.133 | .202 | 141.299 | 1.000 | -.674 | .407 |
| | | 3 | -.562 | .210 | 142.300 | .050 | -1.124 | .000 |
| | 3 | Baseline | .768* | .179 | 139.158 | .000 | .289 | 1.247 |
| | | 1 | .429 | .191 | 141.718 | .157 | -.082 | .939 |
| | | 2 | .562 | .210 | 142.300 | .050 | .000 | 1.124 |
| high | Baseline | 1 | .003 | .200 | 133.424 | 1.000 | -.531 | .538 |
| | | 2 | -.046 | .193 | 137.351 | 1.000 | -.563 | .471 |
| | | 3 | .152 | .236 | 144.385 | 1.000 | -.478 | .782 |
| | 1 | Baseline | -.003 | .200 | 133.424 | 1.000 | -.538 | .531 |
| | | 2 | -.049 | .239 | 152.928 | 1.000 | -.689 | .590 |
| | | 3 | .148 | .267 | 152.724 | 1.000 | -.566 | .863 |

| | | | | | | | | |
|---|---|---|---|---|---|---|---|---|
| | 2 | Baseline | .046 | .193 | 137.351 | 1.000 | -.471 | .563 |
| | | 1 | .049 | .239 | 152.928 | 1.000 | -.590 | .689 |
| | | 3 | .198 | .270 | 156.706 | 1.000 | -.524 | .919 |
| | 3 | Baseline | -.152 | .236 | 144.385 | 1.000 | -.782 | .478 |
| | | 1 | -.148 | .267 | 152.724 | 1.000 | -.863 | .566 |
| | | 2 | -.198 | .270 | 156.706 | 1.000 | -.919 | .524 |

**Table 6: Results mixed model 2 "CERAD-Plus: Trail Making Test – Part A"**

| Tests of Fixed Effects | | | | |
|---|---|---|---|---|
| Source | Num. df | Den. df | F | Sig. |
| Intercept | 1 | 70.374 | 9.833 | .003 |
| TIMECATEGORY | 3 | 135.552 | 6.450 | .000 |
| MEL | 1 | 82.731 | .002 | .966 |
| DaT | 1 | 70.826 | 1.417 | .238 |
| COGNITIONGROUP | 2 | 70.665 | 29.687 | .000 |
| TIMECATEGORY*COGNITIONGROUP | 6 | 135.143 | 4.185 | .001 |

| Estimates (COGNITIONGROUP*TIMECATEGORY) | | | | | | |
|---|---|---|---|---|---|---|
| | | | | | 95 % CI | |
| COGNITIONGROUP | TIMECATEGORY | Mean | Std. Error | df | Lower Bound | Upper Bound |
| low | Baseline | -1.246 | .196 | 131.667 | -1.633 | -.858 |
| | 1 | -.436 | .227 | 160.706 | -.884 | .011 |
| | 2 | -.127 | .261 | 174.520 | -.641 | .387 |
| | 3 | -.224 | .261 | 171.192 | -.739 | .291 |
| medium | Baseline | .171 | .192 | 131.015 | -.209 | .552 |
| | 1 | .778 | .247 | 172.826 | .291 | 1.265 |
| | 2 | .886 | .223 | 157.268 | .445 | 1.327 |
| | 3 | .430 | .222 | 151.411 | -.009 | .868 |
| high | Baseline | 1.354 | .187 | 129.710 | .985 | 1.724 |
| | 1 | 1.371 | .225 | 164.941 | .928 | 1.814 |
| | 2 | .992 | .238 | 171.210 | .523 | 1.461 |
| | 3 | 1.238 | .288 | 178.590 | .669 | 1.806 |

| Pairwise Comparisons (COGNITIONGROUP*TIMECATEGORY) | | | | | | | | |
|---|---|---|---|---|---|---|---|---|
| | | | | | | | 95% CI | |
| COGNITIONGROUP | (I) TIME-CATEGORY | (J) TIME-CATEGORY | Mean Difference (I-J) | Std. Error | df | Sig. | Lower Bound | Upper Bound |
| low | Baseline | 1 | -.809* | .235 | 125.689 | .005 | -1.440 | -.179 |
| | | 2 | -1.119* | .268 | 132.683 | .000 | -1.838 | -.400 |
| | | 3 | -1.022* | .268 | 138.780 | .001 | -1.740 | -.303 |
| | 1 | Baseline | .809* | .235 | 125.689 | .005 | .179 | 1.440 |
| | | 2 | -.310 | .293 | 137.130 | 1.000 | -1.095 | .476 |
| | | 3 | -.212 | .301 | 148.128 | 1.000 | -1.017 | .592 |
| | 2 | Baseline | 1.119* | .268 | 132.683 | .000 | .400 | 1.838 |
| | | 1 | .310 | .293 | 137.130 | 1.000 | -.476 | 1.095 |
| | | 3 | .097 | .324 | 146.374 | 1.000 | -.768 | .963 |
| | 3 | Baseline | 1.022* | .268 | 138.780 | .001 | .303 | 1.740 |
| | | 1 | .212 | .301 | 148.128 | 1.000 | -.592 | 1.017 |
| | | 2 | -.097 | .324 | 146.374 | 1.000 | -.963 | .768 |
| medium | Baseline | 1 | -.607 | .255 | 129.343 | .112 | -1.289 | .076 |
| | | 2 | -.715* | .230 | 129.493 | .014 | -1.331 | -.099 |
| | | 3 | -.258 | .230 | 133.890 | 1.000 | -.874 | .358 |
| | 1 | Baseline | .607 | .255 | 129.343 | .112 | -.076 | 1.289 |
| | | 2 | -.108 | .291 | 145.624 | 1.000 | -.887 | .671 |
| | | 3 | .349 | .284 | 144.304 | 1.000 | -.411 | 1.108 |
| | 2 | Baseline | .715* | .230 | 129.493 | .014 | .099 | 1.331 |
| | | 1 | .108 | .291 | 145.624 | 1.000 | -.671 | .887 |
| | | 3 | .457 | .254 | 137.054 | .450 | -.225 | 1.138 |
| | 3 | Baseline | .258 | .230 | 133.890 | 1.000 | -.358 | .874 |
| | | 1 | -.349 | .284 | 144.304 | 1.000 | -1.108 | .411 |
| | | 2 | -.457 | .254 | 137.054 | .450 | -1.138 | .225 |
| high | Baseline | 1 | -.016 | .233 | 127.575 | 1.000 | -.641 | .608 |

| | | | | | | | | |
|---|---|---|---|---|---|---|---|---|
| | | 2 | .362 | .239 | 129.167 | .794 | -.279 | 1.003 |
| | | 3 | .117 | .295 | 140.704 | 1.000 | -.672 | .906 |
| | 1 | Baseline | .016 | .233 | 127.575 | 1.000 | -.608 | .641 |
| | | 2 | .379 | .282 | 140.462 | 1.000 | -.376 | 1.133 |
| | | 3 | .133 | .320 | 144.146 | 1.000 | -.723 | .990 |
| | 2 | Baseline | -.362 | .239 | 129.167 | .794 | -1.003 | .279 |
| | | 1 | -.379 | .282 | 140.462 | 1.000 | -1.133 | .376 |
| | | 3 | -.245 | .333 | 145.510 | 1.000 | -1.137 | .646 |
| | 3 | Baseline | -.117 | .295 | 140.704 | 1.000 | -.906 | .672 |
| | | 1 | -.133 | .320 | 144.146 | 1.000 | -.990 | .723 |
| | | 2 | .245 | .333 | 145.510 | 1.000 | -.646 | 1.137 |

#### 2.3 UPDRS-III

##### 2.3.1 Statistical analysis

###### 2.3.1.1 Implementation

The UPDRS-III was performed at least once after six months of melatonin treatment with an average follow up of 3.5 ± 2.9 years (range: 0.6-21.7 yrs) in 153/167 iRBD patients. For further analyses, only those patients who were untreated at baseline and examined motorically multiple times (n=75) were included, with 140 follow-up examinations compared at three different time points (=TIMECATEGORY) to motor performance at baseline.

###### 2.3.1.2 Model specification

In model 1, analyses were performed for the UPDRS-III total score as a dependent variable, treating TIMECATEGORY as a fixed, patient ID (=ID) as a random factor and continuity of melatonin intake (=MEL; “continuously” vs. “discontinuously”) as well as DaTSPECT results (=DaT; “normal” vs. “borderline” vs. “pathological” scan) as covariates.
Within model 2, a group factor (MOTORGROUP) was added, categorizing patients into "low", "medium" or "high" performer depending on their motor performance at baseline (“high” performer representing the clinically most impaired patients). Also, the interaction term of TIMECATEGORY*MOTORGROUP was included as an additional fixed factor besides the variables defined in model 1.

###### 2.3.1.3 Descriptive statistics

Means and standard deviations with ranges in years of the follow up duration for the total and sub-cohort as well as the definition of TIMECATEGORIES, the number of re-tests and mean z-scores with ranges of the different MOTORGROUPS are characterized as follows:

**Table 7: Descriptive statistics “UPDRS-III”**

| Follow-up duration and re-examinations | | | Linear mixed model | | | | | |
|---|---|---|---|---|---|---|---|---|
| | | | | | | MOTORGROUP | | |
| Follow up of total cohort (yrs) | Follow up of included sub-cohort (yrs) | Number of re-tests | TIME C. 1 (yrs) | TIME C. 2 (yrs) | TIME C. 3 (yrs) | low | medium | high |
| 3.5 ± 2.9<br>(0.6-21.7)<br>n= 153/167 | 2.3 ± 1.5<br>(0.5-9.3)<br>n= 75/167 | 1x =28<br>2x = 31<br>3x = 14<br>4x = 2<br>n= 140 | 0.9<br>(0.5-1.1)<br>n= 46 | 1.7<br>(1.2-2.3)<br>n= 47 | 3.5<br>(2.4-9.3)<br>n= 47 | 0<br>(0-0)<br>n=30 | 1.5<br>(0.5-2)<br>n=22 | 5.9<br>(3-20)<br>n=23 |

##### 2.3.2 Results model 1 (UPDRS-III)

**Table 8: Results mixed model 1 "UPDRS-III"**

| Tests of Fixed Effects | | | | |
|---|---|---|---|---|
| Source | Num. df | Den. df | F | Sig. |
| Intercept | 1 | 51.729 | 2.838 | .098 |
| TIMECATEGORY | 3 | 104.045 | 9.806 | .000 |
| MEL | 1 | 51.395 | .058 | .811 |
| DaT | 1 | 54.562 | .332 | .567 |

| Estimates | | | | | |
|---|---|---|---|---|---|
| | | | | 95% CI | |
| TIME-CATEGORY | Mean | Std. Error | df | Lower Bound | Upper Bound |
| Baseline | 2.016 | .376 | 66.361 | 1.264 | 2.767 |
| 1 | 1.148 | .403 | 83.705 | .346 | 1.951 |
| 2 | .866 | .408 | 86.896 | .055 | 1.677 |
| 3 | .667 | .427 | 97.817 | -.181 | 1.515 |

| Pairwise Comparisons | | | | | | | |
|---|---|---|---|---|---|---|---|
| | | | | | | 95% CI | |
| (I) TIME-CATEGORY | (J) TIME-CATEGORY | Mean Difference (I-J) | Std. Error | df | Sig. | Lower Bound | Upper Bound |
| Baseline | 1 | .868* | .276 | 102.086 | .013 | .125 | 1.610 |
| | 2 | 1.150* | .282 | 101.899 | .001 | .391 | 1.909 |
| | 3 | 1.349* | .309 | 105.246 | .000 | .517 | 2.181 |
| 1 | Baseline | -.868* | .276 | 102.086 | .013 | -1.610 | -.125 |
| | 2 | .282 | .327 | 104.758 | 1.000 | -.597 | 1.162 |
| | 3 | .481 | .359 | 107.997 | 1.000 | -.482 | 1.445 |
| 2 | Baseline | -1.150* | .282 | 101.899 | .001 | -1.909 | -.391 |
| | 1 | -.282 | .327 | 104.758 | 1.000 | -1.162 | .597 |
| | 3 | .199 | .351 | 105.778 | 1.000 | -.744 | 1.143 |
| 3 | Baseline | -1.349* | .309 | 105.246 | .000 | -2.181 | -.517 |
| | 1 | -.481 | .359 | 107.997 | 1.000 | -1.445 | .482 |
| | 2 | -.199 | .351 | 105.778 | 1.000 | -1.143 | .744 |

##### 2.3.3 Results model 2 (UPDRS-III)

**Table 9: Results mixed model 2 "UPDRS-III"**

| Tests of Fixed Effects | | | | |
|---|---|---|---|---|
| Source | Num. df | Den. Df | F | Sig. |
| Intercept | 1 | 51.122 | 8.396 | .006 |
| TIMECATEGORY | 3 | 103.881 | 28.067 | .000 |
| MEL | 1 | 50.905 | .153 | .697 |
| DaT | 1 | 54.087 | .001 | .979 |
| MOTORGROUP | 2 | 51.436 | 17.146 | .000 |
| TIMECATEGORY*MOTORGROUP | 6 | 103.675 | 13.327 | .000 |

| Estimates (MOTORGROUP*TIMECATEGORY) | | | | | | |
|---|---|---|---|---|---|---|
| MOTOR-GROUP | TIME- CATEGORY | Mean | Std. Error | df | 95 % CI | |
| | | | | | Lower Bound | Upper Bound |
| low | Baseline | -.033 | .430 | 65.446 | -.892 | .826 |
| | 1 | .066 | .457 | 80.371 | -.843 | .975 |
| | 2 | -.010 | .472 | 88.970 | -.948 | .928 |
| | 3 | .169 | .517 | 111.980 | -.855 | 1.193 |
| medium | Baseline | 1.569 | .484 | 65.506 | .602 | 2.536 |
| | 1 | .692 | .518 | 82.291 | -.339 | 1.724 |
| | 2 | .454 | .507 | 77.039 | -.556 | 1.464 |
| | 3 | .465 | .531 | 87.903 | -.591 | 1.521 |
| high | Baseline | 6.571 | .579 | 66.333 | 5.415 | 7.728 |
| | 1 | 3.914 | .639 | 91.114 | 2.645 | 5.182 |
| | 2 | 2.889 | .659 | 99.660 | 1.581 | 4.197 |
| | 3 | 2.334 | .642 | 90.887 | 1.059 | 3.610 |

| Pairwise Comparisons (TIMECATEGORY*MOTORGROUP) | | | | | | | | |
|---|---|---|---|---|---|---|---|---|
| MOTOR-GROUP | (I) TIME-CATEGORY | (J) TIME-CATEGORY | Mean Difference (I-J) | Std. Error | df | Sig. | 95% CI | |
| | | | | | | | Lower Bound | Upper Bound |
| low | Baseline | 1 | -.099 | .311 | 101.164 | 1.000 | -.936 | .738 |
| | | 2 | -.023 | .329 | 102.000 | 1.000 | -.908 | .862 |
| | | 3 | -.202 | .392 | 105.009 | 1.000 | -1.256 | .852 |
| | 1 | Baseline | .099 | .311 | 101.164 | 1.000 | -.738 | .936 |
| | | 2 | .076 | .375 | 104.199 | 1.000 | -.933 | 1.085 |
| | | 3 | -.103 | .438 | 107.599 | 1.000 | -1.280 | 1.074 |
| | 2 | Baseline | .023 | .329 | 102.000 | 1.000 | -.862 | .908 |
| | | 1 | -.076 | .375 | 104.199 | 1.000 | -1.085 | .933 |
| | | 3 | -.179 | .451 | 106.941 | 1.000 | -1.391 | 1.033 |
| | 3 | Baseline | .202 | .392 | 105.009 | 1.000 | -.852 | 1.256 |
| | | 1 | .103 | .438 | 107.599 | 1.000 | -1.074 | 1.280 |
| | | 2 | .179 | .451 | 106.941 | 1.000 | -1.033 | 1.391 |
| medium | Baseline | 1 | .877 | .351 | 101.412 | .085 | -.068 | 1.821 |
| | | 2 | 1.115* | .339 | 99.842 | .008 | .202 | 2.028 |
| | | 3 | 1.104* | .372 | 103.957 | .022 | .103 | 2.105 |
| | 1 | Baseline | -.877 | .351 | 101.412 | .085 | -1.821 | .068 |
| | | 2 | .238 | .392 | 102.922 | 1.000 | -.817 | 1.293 |
| | | 3 | .227 | .437 | 107.286 | 1.000 | -.947 | 1.402 |
| | 2 | Baseline | -1.115* | .339 | 99.842 | .008 | -2.028 | -.202 |
| | | 1 | -.238 | .392 | 102.922 | 1.000 | -1.293 | .817 |
| | | 3 | -.011 | .406 | 103.970 | 1.000 | -1.103 | 1.082 |
| | 3 | Baseline | -1.104* | .372 | 103.957 | .022 | -2.105 | -.103 |
| | | 1 | -.227 | .437 | 107.286 | 1.000 | -1.402 | .947 |
| | | 2 | .011 | .406 | 103.970 | 1.000 | -1.082 | 1.103 |
| high | Baseline | 1 | 2.658* | .457 | 102.005 | .000 | 1.429 | 3.886 |
| | | 2 | 3.683* | .486 | 102.440 | .000 | 2.375 | 4.990 |
| | | 3 | 4.237* | .462 | 105.554 | .000 | 2.995 | 5.479 |
| | 1 | Baseline | -2.658* | .457 | 102.005 | .000 | -3.886 | -1.429 |
| | | 2 | 1.025 | .586 | 107.001 | .501 | -.552 | 2.601 |
| | | 3 | 1.579* | .562 | 108.830 | .035 | .068 | 3.090 |
| | 2 | Baseline | -3.683* | .486 | 102.440 | .000 | -4.990 | -2.375 |
| | | 1 | -1.025 | .586 | 107.001 | .501 | -2.601 | .552 |

| | | | | | | | |
|---|---|---|---|---|---|---|---|
| | | 3 | .554 | .550 | 104.989 | 1.000 | -.924 | 2.033 |
| | 3 | Baseline | -4.237* | .462 | 105.554 | .000 | -5.479 | -2.995 |
| | | 1 | -1.579* | .562 | 108.830 | .035 | -3.090 | -.068 |
| | | 2 | -.554 | .550 | 104.989 | 1.000 | -2.033 | .924 |

#### 2.4 Ikelos-RS/CGI

##### 2.4.1 Validation

To test convergent validity, the Ikelos-RS total score was correlated with the CGI-S in 1153 cases. For the evaluation of the interrater reliability, 45 Ikelos-RS were filled out by two independent physicians. 174 Ikelos-RS were filled out again by the same physician after at least six months. Overall, correlation analyses (using Spearman's rho) showed that the Ikelos-RS can be considered as a valid measuring instrument: Results provide evidence for convergent validity ($\rho$=.9, p<.001), interrater reliability ($\rho$= .9, p<.001) and test-retest reliability ($\rho$= .9, p<.001).

##### 2.4.2 Statistical analysis

###### 2.4.2.1 Implementation

RBD symptom severity was evaluated with a total of 1489 Ikelos-RS at the time of diagnosis and each further visit (usually at least three times per year since 2016) after initiation of treatment, with an average follow up of 4.2±3.1 years (range: 0.6-21.7 years) in all 167 patients under longterm chronobiotic treatment and with an average follow up of 3.1 ± 2.1 years (range: 0.3-7.6 years) in 39 patients under mixed treatments.

To investigate the influence of confounding factors on RBD symptom regression within the first 6 months of melatonin treatment, time courses of the following four subgroups in the group under longterm chronobiotic treatment were compared: 1) patients with beta blockers (n=41), 2) with antidepressants (n=18), 3) with inadequate timing (n=25) and 4) without such confounders (n=77). Because no clear subgroup classification could be determined, data of patients who took both beta blockers and antidepressants (n=6) were not included in the analysis of the first 6 months (model A). For the group under mixed treatments two subgroups were compared: 1) patients short term (up to 3 months) treated with melatonin (n=13), and 2) patients treated with other medications (n=26).

To investigate differences in time courses after six months, the patients of the longterm chronobiotic treatment group were re-classified in patients who 1) continued to take melatonin regularly (n=137) and 2) stopped after 6 months at the latest (n=30), thus including all 167 patients (model B, I). Additionally, time courses of these two groups were compared to both sub-classifications of the patients under mixed treatments (model B, II).

###### 2.4.2.2 Model specification

A random intercept model was performed, treating the Ikelos-RS total score as a dependent variable (=IKELOS). Due to a non-symmetrical distribution, the time variable (months since melatonin treatment) was both logarithmized and squared. Subsequently, time (=ln_TIME) and subgroup classification (=GROUP) were defined as fixed and patient as random factors (=ID). Additionally, the interaction term of ln_TIME*GROUP was included.

##### 2.4.3 Results model A (Ikelos-RS)

**Table 10: Results mixed model A "Ikelos-RS"**

| Tests of Fixed Effects | | | | |
|---|---|---|---|---|
| Source | Num. df | Den. Df | F | Sig. |
| Intercept | 1 | 913.942 | 3161.420 | 0.000 |
| GROUP | 5 | 893.329 | 5.463 | 0.000 |
| ln_TIME | 1 | 1411.655 | 398.375 | 0.000 |
| ln_TIME* ln_TIME | 1 | 1438.178 | 178.362 | 0.000 |
| GROUP* ln_TIME | 5 | 1413.507 | 12.259 | 0.000 |
| GROUP* ln_TIME* ln_TIME | 5 | 1420.760 | 10.245 | 0.000 |

| Estimates of Fixed Effects | | | | | | | |
|---|---|---|---|---|---|---|---|
| | | | | | | 95 % CI | |
| Parameter | Estimate | Std. Error | df | t | Sig. | Lower Bound | Upper Bound |
| Intercept | 5.184824 | 0.132887 | 833.039 | 39.017 | 0.000 | 4.923992 | 5.445657 |
| GROUP (reference: no confounder) | | | | | | | |
| GROUP (timing) | 1.139871 | 0.272585 | 861.276 | 4.182 | 0.000 | 0.604863 | 1.674880 |
| GROUP (beta blockers) | 0.275746 | 0.227301 | 836.604 | 1.213 | 0.225 | -0.170400 | 0.721892 |
| GROUP (antidepressants) | -0.068688 | 0.309461 | 855.274 | -0.222 | 0.824 | -0.676079 | 0.538703 |
| GROUP (short term melatonin) | 0.665816 | 0.373709 | 967.846 | 1.782 | 0.075 | -0.067556 | 1.399189 |
| GROUP (various) | 0.951135 | 0.276533 | 921.235 | 3.440 | 0.001 | 0.408428 | 1.493842 |
| ln_TIME | -2.568642 | 0.124480 | 1386.165 | -20.635 | 0.000 | -2.812832 | -2.324452 |
| ln_TIME* ln_TIME | 0.408138 | 0.028640 | 1429.236 | 14.251 | 0.000 | 0.351958 | 0.464319 |
| GROUP* ln_TIME (reference: no confounder) | | | | | | | |
| GROUP (timing) * ln_TIME | 0.824670 | 0.250492 | 1381.328 | 3.292 | 0.001 | 0.333284 | 1.316056 |
| GROUP (beta blockers) * ln_TIME | -0.066039 | 0.206288 | 1443.941 | -0.320 | 0.749 | -0.470695 | 0.338617 |
| GROUP (antidepressants) * ln_TIME | 0.439222 | 0.297827 | 1430.603 | 1.475 | 0.140 | -0.145003 | 1.023446 |
| GROUP (short term melatonin) * ln_TIME | -0.744790 | 0.434854 | 1371.218 | -1.713 | 0.087 | -1.597841 | 0.108260 |
| GROUP (various) * ln_TIME | 2.060543 | 0.310628 | 1426.684 | 6.633 | 0.000 | 1.451205 | 2.669880 |
| GROUP* ln_TIME* ln_TIME (reference: no confounder) | | | | | | | |
| GROUP (timing) * ln_TIME* ln_TIME | -0.256591 | 0.056744 | 1423.681 | -4.522 | 0.000 | -0.367901 | -0.145281 |
| GROUP (beta blockers) * ln_TIME* ln_TIME | -0.008078 | 0.047190 | 1392.294 | -0.171 | 0.864 | -0.100648 | 0.084493 |
| GROUP (antidepressants) * ln_TIME* ln_TIME | -0.115238 | 0.065884 | 1443.924 | -1.749 | 0.080 | -0.244476 | 0.014001 |
| GROUP (short term melatonin) * ln_TIME* ln_TIME | 0.323664 | 0.109873 | 1408.042 | 2.946 | 0.003 | 0.108131 | 0.539197 |
| GROUP (various) * ln_TIME* ln_TIME | -0.322552 | 0.073796 | 1443.996 | -4.371 | 0.000 | -0.467311 | -0.177793 |

| Estimates (Baseline) | | | | | |
|---|---|---|---|---|---|
| | | | | 95 % CI | |
| GROUP | Mean | Std. Error | df | Lower Bound | Upper Bound |
| no confounder | 5.185 | 0.133 | 833.039 | 4.924 | 5.446 |
| timing | 6.325 | 0.238 | 870.229 | 5.858 | 6.792 |
| beta blockers | 5.461 | 0.184 | 838.459 | 5.099 | 5.823 |
| antidepressants | 5.116 | 0.279 | 860.365 | 4.568 | 5.665 |
| short term melatonin | 5.851 | 0.349 | 988.360 | 5.165 | 6.536 |
| various | 6.136 | 0.243 | 948.887 | 5.660 | 6.612 |

| Pairwise Comparisons (Baseline) | | | | | | | |
|---|---|---|---|---|---|---|---|
| | | | | | | 95 % CI | |
| (I)GROUP | (J)GROUP | Mean Difference (I-J) | Std. Error | df | Sig. | Lower Bound | Upper Bound |
| no confounder | timing | -1.140* | 0.273 | 861.276 | 0.000 | -1.942 | -0.338 |
| | beta blockers | -0.276 | 0.227 | 836.604 | 1.000 | -0.945 | 0.393 |
| | antidepressants | 0.069 | 0.309 | 855.274 | 1.000 | -0.842 | 0.980 |
| | short term | -0.666 | 0.374 | 967.846 | 1.000 | -1.765 | 0.434 |
| | various | -.951* | 0.277 | 921.235 | 0.009 | -1.765 | -0.137 |
| timing | beta blockers | 0.864 | 0.301 | 858.204 | 0.063 | -0.022 | 1.750 |
| | antidepressants | 1.209* | 0.367 | 864.501 | 0.016 | 0.128 | 2.289 |
| | short term | 0.474 | 0.423 | 949.963 | 1.000 | -0.770 | 1.718 |
| | various | 0.189 | 0.340 | 909.745 | 1.000 | -0.811 | 1.189 |
| | no confounder | 1.140* | 0.273 | 861.276 | 0.000 | 0.338 | 1.942 |
| beta blockers | timing | -0.864 | 0.301 | 858.204 | 0.063 | -1.750 | 0.022 |
| | antidepressants | 0.344 | 0.335 | 853.674 | 1.000 | -0.641 | 1.330 |

| | | | | | | | |
|---|---|---|---|---|---|---|---|
| | short term | -0.390 | 0.395 | 954.416 | 1.000 | -1.552 | 0.772 |
| | various | -0.675 | 0.305 | 907.354 | 0.403 | -1.572 | 0.221 |
| | no confounder | 0.276 | 0.227 | 836.604 | 1.000 | -0.393 | 0.945 |
| antidepressants | timing | -1.209* | 0.367 | 864.501 | 0.016 | -2.289 | -0.128 |
| | beta blockers | -0.344 | 0.335 | 853.674 | 1.000 | -1.330 | 0.641 |
| | short term | -0.735 | 0.447 | 937.136 | 1.000 | -2.051 | 0.582 |
| | various | -1.020 | 0.370 | 897.681 | 0.090 | -2.109 | 0.069 |
| | no confounder | -0.069 | 0.309 | 855.274 | 1.000 | -0.980 | 0.842 |
| short term | timing | -0.474 | 0.423 | 949.963 | 1.000 | -1.718 | 0.770 |
| | beta blockers | 0.390 | 0.395 | 954.416 | 1.000 | -0.772 | 1.552 |
| | antidepressants | 0.735 | 0.447 | 937.136 | 1.000 | -0.582 | 2.051 |
| | various | -0.285 | 0.425 | 975.437 | 1.000 | -1.537 | 0.966 |
| | no confounder | 0.666 | 0.374 | 967.846 | 1.000 | -0.434 | 1.765 |
| various | timing | -0.189 | 0.340 | 909.745 | 1.000 | -1.189 | 0.811 |
| | beta blockers | 0.675 | 0.305 | 907.354 | 0.403 | -0.221 | 1.572 |
| | antidepressants | 1.020 | 0.370 | 897.681 | 0.090 | -0.069 | 2.109 |
| | short term | 0.285 | 0.425 | 975.437 | 1.000 | -0.966 | 1.537 |
| | no confounder | .951* | 0.277 | 921.235 | 0.009 | 0.137 | 1.765 |

**Estimates (2 weeks)**

| GROUP | Mean | Std. Error | df | 95 % CI Lower Bound | 95 % CI Upper Bound |
|---|---|---|---|---|---|
| no confounder | 4.268 | 0.110 | 476.873 | 4.051 | 4.484 |
| timing | 5.684 | 0.196 | 500.616 | 5.299 | 6.069 |
| beta blockers | 4.517 | 0.153 | 504.510 | 4.216 | 4.819 |
| antidepressants | 4.349 | 0.232 | 497.518 | 3.893 | 4.805 |
| short term melatonin | 4.697 | 0.283 | 574.907 | 4.140 | 5.254 |
| various | 5.955 | 0.199 | 553.495 | 5.564 | 6.346 |

**Pairwise Comparisons (2 weeks)**

| (I)GROUP | (J)GROUP | Mean Difference (I-J) | Std. Error | df | Sig. | 95 % CI Lower Bound | 95 % CI Upper Bound |
|---|---|---|---|---|---|---|---|
| no confounder | timing | -1.416* | 0.225 | 494.770 | 0.000 | -2.079 | -0.753 |
| | beta blockers | -0.249 | 0.189 | 494.881 | 1.000 | -0.807 | 0.308 |
| | antidepressants | -0.082 | 0.257 | 493.634 | 1.000 | -0.839 | 0.676 |
| | short term | -0.430 | 0.304 | 560.722 | 1.000 | -1.326 | 0.467 |
| | various | -1.688* | 0.228 | 534.244 | 0.000 | -2.359 | -1.016 |
| timing | beta blockers | 1.167* | 0.249 | 502.093 | 0.000 | 0.433 | 1.901 |
| | antidepressants | 1.335* | 0.304 | 498.804 | 0.000 | 0.439 | 2.230 |
| | short term | 0.987 | 0.345 | 549.537 | 0.065 | -0.029 | 2.002 |
| | various | -0.271 | 0.279 | 526.690 | 1.000 | -1.095 | 0.552 |
| | no confounder | 1.416* | 0.225 | 494.770 | 0.000 | 0.753 | 2.079 |
| beta blockers | timing | -1.167* | 0.249 | 502.093 | 0.000 | -1.901 | -0.433 |
| | antidepressants | 0.168 | 0.278 | 499.632 | 1.000 | -0.653 | 0.989 |
| | short term | -0.180 | 0.322 | 557.972 | 1.000 | -1.130 | 0.770 |
| | various | -1.438* | 0.251 | 534.600 | 0.000 | -2.179 | -0.697 |
| | no confounder | 0.249 | 0.189 | 494.881 | 1.000 | -0.308 | 0.807 |
| antidepressants | timing | -1.335* | 0.304 | 498.804 | 0.000 | -2.230 | -0.439 |
| | beta blockers | -0.168 | 0.278 | 499.632 | 1.000 | -0.989 | 0.653 |
| | short term | -0.348 | 0.366 | 542.220 | 1.000 | -1.428 | 0.732 |
| | various | -1.606* | 0.306 | 520.371 | 0.000 | -2.508 | -0.704 |
| | no confounder | 0.082 | 0.257 | 493.634 | 1.000 | -0.676 | 0.839 |
| short term | timing | -0.987 | 0.345 | 549.537 | 0.065 | -2.002 | 0.029 |
| | beta blockers | 0.180 | 0.322 | 557.972 | 1.000 | -0.770 | 1.130 |
| | antidepressants | 0.348 | 0.366 | 542.220 | 1.000 | -0.732 | 1.428 |

| | | | | | | | |
|---|---|---|---|---|---|---|---|
| | various | -1.258* | 0.346 | 567.725 | 0.005 | -2.279 | -0.237 |
| | no confounder | 0.430 | 0.304 | 560.722 | 1.000 | -0.467 | 1.326 |
| various | timing | 0.271 | 0.279 | 526.690 | 1.000 | -0.552 | 1.095 |
| | beta blockers | 1.438* | 0.251 | 534.600 | 0.000 | 0.697 | 2.179 |
| | antidepressants | 1.606* | 0.306 | 520.371 | 0.000 | 0.704 | 2.508 |
| | short term | 1.258* | 0.346 | 567.725 | 0.005 | 0.237 | 2.279 |
| | no confounder | 1.688* | 0.228 | 534.244 | 0.000 | 1.016 | 2.359 |

**Estimates (4 weeks)**

| GROUP | Mean | Std. Error | df | 95 % CI | |
|---|---|---|---|---|---|
| | | | | Lower Bound | Upper Bound |
| no confounder | 3.688 | 0.102 | 357.151 | 3.486 | 3.889 |
| timing | 5.255 | 0.180 | 365.650 | 4.901 | 5.609 |
| beta blockers | 3.917 | 0.142 | 366.008 | 3.638 | 4.196 |
| antidepressants | 3.856 | 0.217 | 374.218 | 3.429 | 4.283 |
| short term melatonin | 4.006 | 0.271 | 481.474 | 3.473 | 4.539 |
| various | 5.842 | 0.192 | 456.678 | 5.465 | 6.219 |

**Pairwise Comparisons (4 weeks)**

| (I)GROUP | (J)GROUP | Mean Difference (I-J) | Std. Error | df | Sig. | 95 % CI | |
|---|---|---|---|---|---|---|---|
| | | | | | | Lower Bound | Upper Bound |
| no confounder | timing | -1.567* | 0.207 | 363.546 | 0.000 | -2.179 | -0.956 |
| | beta blockers | -0.229 | 0.175 | 362.937 | 1.000 | -0.746 | 0.287 |
| | antidepressants | -0.168 | 0.240 | 371.036 | 1.000 | -0.878 | 0.541 |
| | short term | -0.318 | 0.290 | 463.199 | 1.000 | -1.174 | 0.537 |
| | various | -2.154* | 0.217 | 431.774 | 0.000 | -2.796 | -1.512 |
| timing | beta blockers | 1.338* | 0.229 | 365.787 | 0.000 | 0.661 | 2.015 |
| | antidepressants | 1.399* | 0.282 | 370.699 | 0.000 | 0.566 | 2.233 |
| | short term | 1.249* | 0.325 | 441.596 | 0.002 | 0.289 | 2.210 |
| | various | -0.587 | 0.263 | 410.794 | 0.394 | -1.363 | 0.190 |
| | no confounder | 1.567* | 0.207 | 363.546 | 0.000 | 0.956 | 2.179 |
| beta blockers | timing | -1.338* | 0.229 | 365.787 | 0.000 | -2.015 | -0.661 |
| | antidepressants | 0.061 | 0.259 | 371.742 | 1.000 | -0.705 | 0.827 |
| | short term | -0.089 | 0.306 | 453.141 | 1.000 | -0.992 | 0.814 |
| | various | -1.925* | 0.239 | 421.649 | 0.000 | -2.629 | -1.221 |
| | no confounder | 0.229 | 0.175 | 362.937 | 1.000 | -0.287 | 0.746 |
| antidepressants | timing | -1.399* | 0.282 | 370.699 | 0.000 | -2.233 | -0.566 |
| | beta blockers | -0.061 | 0.259 | 371.742 | 1.000 | -0.827 | 0.705 |
| | short term | -0.150 | 0.347 | 435.361 | 1.000 | -1.176 | 0.875 |
| | various | -1.986* | 0.290 | 407.750 | 0.000 | -2.842 | -1.130 |
| | no confounder | 0.168 | 0.240 | 371.036 | 1.000 | -0.541 | 0.878 |
| short term | timing | -1.249* | 0.325 | 441.596 | 0.002 | -2.210 | -0.289 |
| | beta blockers | 0.089 | 0.306 | 453.141 | 1.000 | -0.814 | 0.992 |
| | antidepressants | 0.150 | 0.347 | 435.361 | 1.000 | -0.875 | 1.176 |
| | various | -1.836* | 0.332 | 473.019 | 0.000 | -2.816 | -0.856 |
| | no confounder | 0.318 | 0.290 | 463.199 | 1.000 | -0.537 | 1.174 |
| various | timing | 0.587 | 0.263 | 410.794 | 0.394 | -0.190 | 1.363 |
| | beta blockers | 1.925* | 0.239 | 421.649 | 0.000 | 1.221 | 2.629 |
| | antidepressants | 1.986* | 0.290 | 407.750 | 0.000 | 1.130 | 2.842 |
| | short term | 1.836* | 0.332 | 473.019 | 0.000 | 0.856 | 2.816 |
| | no confounder | 2.154* | 0.217 | 431.774 | 0.000 | 1.512 | 2.796 |

| Estimates (6 weeks) | | | | | |
|---|---|---|---|---|---|
| | | | | 95 % CI | |
| GROUP | Mean | Std. Error | df | Lower Bound | Upper Bound |
| no confounder | 3.259 | 0.100 | 318.005 | 3.062 | 3.456 |
| timing | 4.922 | 0.174 | 316.918 | 4.580 | 5.265 |
| beta blockers | 3.471 | 0.137 | 307.938 | 3.201 | 3.741 |
| antidepressants | 3.485 | 0.214 | 336.538 | 3.064 | 3.907 |
| short term melatonin | 3.522 | 0.276 | 493.357 | 2.980 | 4.064 |
| various | 5.759 | 0.196 | 458.801 | 5.373 | 6.144 |

| Pairwise Comparisons (6 weeks) | | | | | | | |
|---|---|---|---|---|---|---|---|
| | | | | | | 95 % CI | |
| (I)GROUP | (J)GROUP | Mean Difference (I-J) | Std. Error | df | Sig. | Lower Bound | Upper Bound |
| no confounder | timing | -1.663* | 0.201 | 317.187 | 0.000 | -2.257 | -1.069 |
| | beta blockers | -0.212 | 0.170 | 311.378 | 1.000 | -0.714 | 0.290 |
| | antidepressants | -0.226 | 0.236 | 333.120 | 1.000 | -0.925 | 0.473 |
| | short term | -0.263 | 0.293 | 467.430 | 1.000 | -1.129 | 0.603 |
| | various | -2.500* | 0.220 | 423.878 | 0.000 | -3.149 | -1.850 |
| timing | beta blockers | 1.451* | 0.222 | 313.435 | 0.000 | 0.795 | 2.107 |
| | antidepressants | 1.437* | 0.276 | 328.544 | 0.000 | 0.620 | 2.253 |
| | short term | 1.400* | 0.326 | 432.261 | 0.000 | 0.437 | 2.363 |
| | various | -.837* | 0.262 | 387.628 | 0.023 | -1.611 | -0.062 |
| | no confounder | 1.663* | 0.201 | 317.187 | 0.000 | 1.069 | 2.257 |
| beta blockers | timing | -1.451* | 0.222 | 313.435 | 0.000 | -2.107 | -0.795 |
| | antidepressants | -0.014 | 0.254 | 327.860 | 1.000 | -0.766 | 0.738 |
| | short term | -0.051 | 0.308 | 446.868 | 1.000 | -0.960 | 0.859 |
| | various | -2.287* | 0.239 | 400.131 | 0.000 | -2.994 | -1.581 |
| | no confounder | 0.212 | 0.170 | 311.378 | 1.000 | -0.290 | 0.714 |
| antidepressants | timing | -1.437* | 0.276 | 328.544 | 0.000 | -2.253 | -0.620 |
| | beta blockers | 0.014 | 0.254 | 327.860 | 1.000 | -0.738 | 0.766 |
| | short term | -0.037 | 0.349 | 425.019 | 1.000 | -1.068 | 0.995 |
| | various | -2.273* | 0.290 | 386.125 | 0.000 | -3.131 | -1.416 |
| | no confounder | 0.226 | 0.236 | 333.120 | 1.000 | -0.473 | 0.925 |
| short term | timing | -1.400* | 0.326 | 432.261 | 0.000 | -2.363 | -0.437 |
| | beta blockers | 0.051 | 0.308 | 446.868 | 1.000 | -0.859 | 0.960 |
| | antidepressants | 0.037 | 0.349 | 425.019 | 1.000 | -0.995 | 1.068 |
| | various | -2.237* | 0.338 | 481.413 | 0.000 | -3.235 | -1.238 |
| | no confounder | 0.263 | 0.293 | 467.430 | 1.000 | -0.603 | 1.129 |
| various | timing | .837* | 0.262 | 387.628 | 0.023 | 0.062 | 1.611 |
| | beta blockers | 2.287* | 0.239 | 400.131 | 0.000 | 1.581 | 2.994 |
| | antidepressants | 2.273* | 0.290 | 386.125 | 0.000 | 1.416 | 3.131 |
| | short term | 2.237* | 0.338 | 481.413 | 0.000 | 1.238 | 3.235 |
| | no confounder | 2.500* | 0.220 | 423.878 | 0.000 | 1.850 | 3.149 |

| Estimates (8 weeks) | | | | | |
|---|---|---|---|---|---|
| | | | | 95 % CI | |
| GROUP | Mean | Std. Error | df | Lower Bound | Upper Bound |
| no confounder | 2.955 | 0.100 | 309.412 | 2.758 | 3.151 |
| timing | 4.675 | 0.173 | 302.765 | 4.335 | 5.015 |
| beta blockers | 3.153 | 0.136 | 285.056 | 2.885 | 3.421 |
| antidepressants | 3.218 | 0.216 | 330.971 | 2.794 | 3.643 |
| short term melatonin | 3.196 | 0.284 | 532.103 | 2.638 | 3.755 |
| various | 5.700 | 0.203 | 487.739 | 5.302 | 6.098 |
| Pairwise Comparisons (8 weeks) | | | | | |

| (I)GROUP | (J)GROUP | Mean Difference (I-J) | Std. Error | df | Sig. | 95 % CI Lower Bound | Upper Bound |
|---|---|---|---|---|---|---|---|
| no confounder | timing | -1.720* | 0.200 | 304.409 | 0.000 | -2.310 | -1.130 |
| | beta blockers | -0.198 | 0.169 | 293.268 | 1.000 | -0.698 | 0.301 |
| | antidepressants | -0.263 | 0.238 | 327.030 | 1.000 | -0.967 | 0.440 |
| | short term | -0.241 | 0.301 | 499.277 | 1.000 | -1.130 | 0.647 |
| | various | -2.745* | 0.226 | 444.000 | 0.000 | -3.412 | -2.079 |
| timing | beta blockers | 1.522* | 0.220 | 295.816 | 0.000 | 0.871 | 2.173 |
| | antidepressants | 1.457* | 0.277 | 319.547 | 0.000 | 0.639 | 2.274 |
| | short term | 1.479* | 0.333 | 452.725 | 0.000 | 0.497 | 2.461 |
| | various | -1.025* | 0.266 | 395.496 | 0.002 | -1.812 | -0.239 |
| | no confounder | 1.720* | 0.200 | 304.409 | 0.000 | 1.130 | 2.310 |
| beta blockers | timing | -1.522* | 0.220 | 295.816 | 0.000 | -2.173 | -0.871 |
| | antidepressants | -0.065 | 0.255 | 316.953 | 1.000 | -0.820 | 0.690 |
| | short term | -0.043 | 0.315 | 469.423 | 1.000 | -0.973 | 0.887 |
| | various | -2.547* | 0.244 | 408.645 | 0.000 | -3.267 | -1.826 |
| | no confounder | 0.198 | 0.169 | 293.268 | 1.000 | -0.301 | 0.698 |
| antidepressants | timing | -1.457* | 0.277 | 319.547 | 0.000 | -2.274 | -0.639 |
| | beta blockers | 0.065 | 0.255 | 316.953 | 1.000 | -0.690 | 0.820 |
| | short term | 0.022 | 0.357 | 443.892 | 1.000 | -1.032 | 1.076 |
| | various | -2.482* | 0.296 | 394.583 | 0.000 | -3.356 | -1.607 |
| | no confounder | 0.263 | 0.238 | 327.030 | 1.000 | -0.440 | 0.967 |
| short term | timing | -1.479* | 0.333 | 452.725 | 0.000 | -2.461 | -0.497 |
| | beta blockers | 0.043 | 0.315 | 469.423 | 1.000 | -0.887 | 0.973 |
| | antidepressants | -0.022 | 0.357 | 443.892 | 1.000 | -1.076 | 1.032 |
| | various | -2.504* | 0.349 | 516.633 | 0.000 | -3.533 | -1.474 |
| | no confounder | 0.241 | 0.301 | 499.277 | 1.000 | -0.647 | 1.130 |
| various | timing | 1.025* | 0.266 | 395.496 | 0.002 | 0.239 | 1.812 |
| | beta blockers | 2.547* | 0.244 | 408.645 | 0.000 | 1.826 | 3.267 |
| | antidepressants | 2.482* | 0.296 | 394.583 | 0.000 | 1.607 | 3.356 |
| | short term | 2.504* | 0.349 | 516.633 | 0.000 | 1.474 | 3.533 |
| | no confounder | 2.745* | 0.226 | 444.000 | 0.000 | 2.079 | 3.412 |

**Estimates (3 months)**

| GROUP | Mean | Std. Error | df | 95 % CI Lower Bound | Upper Bound |
|---|---|---|---|---|---|
| no confounder | 2.505 | 0.101 | 317.089 | 2.306 | 2.705 |
| timing | 4.287 | 0.174 | 304.192 | 3.944 | 4.630 |
| beta blockers | 2.680 | 0.137 | 271.363 | 2.411 | 2.949 |
| antidepressants | 2.816 | 0.222 | 346.000 | 2.378 | 3.253 |
| short term melatonin | 2.752 | 0.301 | 616.372 | 2.161 | 3.343 |
| various | 5.614 | 0.215 | 556.263 | 5.192 | 6.037 |

**Pairwise Comparisons (3 months)**

| (I)GROUP | (J)GROUP | Mean Difference (I-J) | Std. Error | df | Sig. | 95 % CI Lower Bound | Upper Bound |
|---|---|---|---|---|---|---|---|
| no confounder | timing | -1.781* | 0.202 | 307.377 | 0.000 | -2.378 | -1.185 |
| | beta blockers | -0.174 | 0.170 | 286.473 | 1.000 | -0.678 | 0.329 |
| | antidepressants | -0.310 | 0.244 | 340.794 | 1.000 | -1.033 | 0.412 |
| | short term | -0.247 | 0.317 | 573.114 | 1.000 | -1.182 | 0.689 |
| | various | -3.109* | 0.238 | 499.100 | 0.000 | -3.811 | -2.407 |
| timing | beta blockers | 1.607* | 0.221 | 291.090 | 0.000 | 0.951 | 2.262 |
| | antidepressants | 1.471* | 0.283 | 329.231 | 0.000 | 0.635 | 2.307 |
| | short term | 1.535* | 0.348 | 509.593 | 0.000 | 0.510 | 2.560 |

| | | | | | | | |
|---|---|---|---|---|---|---|---|
| | various | -1.328* | 0.277 | 432.385 | 0.000 | -2.145 | -0.510 |
| | no confounder | 1.781* | 0.202 | 307.377 | 0.000 | 1.185 | 2.378 |
| beta blockers | timing | -1.607* | 0.221 | 291.090 | 0.000 | -2.262 | -0.951 |
| | antidepressants | -0.136 | 0.261 | 323.037 | 1.000 | -0.908 | 0.636 |
| | short term | -0.072 | 0.330 | 528.538 | 1.000 | -1.046 | 0.902 |
| | various | -2.935* | 0.255 | 445.386 | 0.000 | -3.687 | -2.182 |
| | no confounder | 0.174 | 0.170 | 286.473 | 1.000 | -0.329 | 0.678 |
| antidepressants | timing | -1.471* | 0.283 | 329.231 | 0.000 | -2.307 | -0.635 |
| | beta blockers | 0.136 | 0.261 | 323.037 | 1.000 | -0.636 | 0.908 |
| | short term | 0.064 | 0.374 | 497.667 | 1.000 | -1.040 | 1.167 |
| | various | -2.799* | 0.310 | 431.873 | 0.000 | -3.712 | -1.885 |
| | no confounder | 0.310 | 0.244 | 340.794 | 1.000 | -0.412 | 1.033 |
| short term | timing | -1.535* | 0.348 | 509.593 | 0.000 | -2.560 | -0.510 |
| | beta blockers | 0.072 | 0.330 | 528.538 | 1.000 | -0.902 | 1.046 |
| | antidepressants | -0.064 | 0.374 | 497.667 | 1.000 | -1.167 | 1.040 |
| | various | -2.862* | 0.370 | 595.206 | 0.000 | -3.952 | -1.772 |
| | no confounder | 0.247 | 0.317 | 573.114 | 1.000 | -0.689 | 1.182 |
| various | timing | 1.328* | 0.277 | 432.385 | 0.000 | 0.510 | 2.145 |
| | beta blockers | 2.935* | 0.255 | 445.386 | 0.000 | 2.182 | 3.687 |
| | antidepressants | 2.799* | 0.310 | 431.873 | 0.000 | 1.885 | 3.712 |
| | short term | 2.862* | 0.370 | 595.206 | 0.000 | 1.772 | 3.952 |
| | no confounder | 3.109* | 0.238 | 499.100 | 0.000 | 2.407 | 3.811 |

**Estimates (4 months)**

| GROUP | Mean | Std. Error | df | 95 % CI Lower Bound | 95 % CI Upper Bound |
|---|---|---|---|---|---|
| no confounder | 2.197 | 0.103 | 330.141 | 1.995 | 2.399 |
| timing | 3.998 | 0.177 | 314.548 | 3.651 | 4.346 |
| beta blockers | 2.352 | 0.138 | 271.058 | 2.081 | 2.623 |
| antidepressants | 2.531 | 0.228 | 366.432 | 2.082 | 2.981 |
| short term melatonin | 2.483 | 0.312 | 678.346 | 1.871 | 3.096 |
| various | 5.556 | 0.224 | 609.720 | 5.116 | 5.997 |

**Pairwise Comparisons (4 months)**

| (I)GROUP | (J)GROUP | Mean Difference (I-J) | Std. Error | df | Sig. | 95 % CI Lower Bound | 95 % CI Upper Bound |
|---|---|---|---|---|---|---|---|
| no confounder | timing | -1.801* | 0.204 | 318.394 | 0.000 | -2.406 | -1.197 |
| | beta blockers | -0.155 | 0.172 | 290.370 | 1.000 | -0.664 | 0.354 |
| | antidepressants | -0.334 | 0.251 | 359.965 | 1.000 | -1.075 | 0.406 |
| | short term | -0.286 | 0.328 | 629.056 | 1.000 | -1.254 | 0.681 |
| | various | -3.359* | 0.247 | 544.321 | 0.000 | -4.086 | -2.633 |
| timing | beta blockers | 1.646* | 0.224 | 297.040 | 0.000 | 0.984 | 2.309 |
| | antidepressants | 1.467* | 0.289 | 345.760 | 0.000 | 0.613 | 2.320 |
| | short term | 1.515* | 0.358 | 555.596 | 0.000 | 0.458 | 2.572 |
| | various | -1.558* | 0.285 | 466.457 | 0.000 | -2.400 | -0.716 |
| | no confounder | 1.801* | 0.204 | 318.394 | 0.000 | 1.197 | 2.406 |
| beta blockers | timing | -1.646* | 0.224 | 297.040 | 0.000 | -2.309 | -0.984 |
| | antidepressants | -0.180 | 0.267 | 337.033 | 1.000 | -0.968 | 0.609 |
| | short term | -0.132 | 0.341 | 575.443 | 1.000 | -1.137 | 0.874 |
| | various | -3.205* | 0.263 | 478.924 | 0.000 | -3.981 | -2.428 |
| | no confounder | 0.155 | 0.172 | 290.370 | 1.000 | -0.354 | 0.664 |
| antidepressants | timing | -1.467* | 0.289 | 345.760 | 0.000 | -2.320 | -0.613 |
| | beta blockers | 0.180 | 0.267 | 337.033 | 1.000 | -0.609 | 0.968 |
| | short term | 0.048 | 0.387 | 541.930 | 1.000 | -1.092 | 1.188 |
| | various | -3.025* | 0.320 | 466.494 | 0.000 | -3.969 | -2.081 |

| | | | | | | | |
|---|---|---|---|---|---|---|---|
| | no confounder | 0.334 | 0.251 | 359.965 | 1.000 | -0.406 | 1.075 |
| short term | timing | -1.515* | 0.358 | 555.596 | 0.000 | -2.572 | -0.458 |
| | beta blockers | 0.132 | 0.341 | 575.443 | 1.000 | -0.874 | 1.137 |
| | antidepressants | -0.048 | 0.387 | 541.930 | 1.000 | -1.188 | 1.092 |
| | various | -3.073* | 0.384 | 654.083 | 0.000 | -4.205 | -1.941 |
| | no confounder | 0.286 | 0.328 | 629.056 | 1.000 | -0.681 | 1.254 |
| various | timing | 1.558* | 0.285 | 466.457 | 0.000 | 0.716 | 2.400 |
| | beta blockers | 3.205* | 0.263 | 478.924 | 0.000 | 2.428 | 3.981 |
| | antidepressants | 3.025* | 0.320 | 466.494 | 0.000 | 2.081 | 3.969 |
| | short term | 3.073* | 0.384 | 654.083 | 0.000 | 1.941 | 4.205 |
| | no confounder | 3.359* | 0.247 | 544.321 | 0.000 | 2.633 | 4.086 |

| **Estimates (6 months)** | | | | | |
|---|---|---|---|---|---|
| | | | | 95 % CI | |
| GROUP | Mean | Std. Error | df | Lower Bound | Upper Bound |
| no confounder | 1.809 | 0.104 | 343.911 | 1.604 | 2.013 |
| timing | 3.593 | 0.179 | 327.502 | 3.241 | 3.946 |
| beta blockers | 1.933 | 0.139 | 274.208 | 1.659 | 2.206 |
| antidepressants | 2.158 | 0.235 | 393.772 | 1.696 | 2.621 |
| short term melatonin | 2.214 | 0.322 | 739.596 | 1.582 | 2.845 |
| various | 5.485 | 0.232 | 667.000 | 5.029 | 5.941 |

| **Pairwise Comparisons (6 months)** | | | | | | | |
|---|---|---|---|---|---|---|---|
| | | | | | | 95 % CI | |
| (I)GROUP | (J)GROUP | Mean Difference (I-J) | Std. Error | df | Sig. | Lower Bound | Upper Bound |
| no confounder | timing | -1.785* | 0.207 | 331.538 | 0.000 | -2.398 | -1.172 |
| | beta blockers | -0.124 | 0.174 | 296.845 | 1.000 | -0.638 | 0.390 |
| | antidepressants | -0.350 | 0.257 | 384.985 | 1.000 | -1.110 | 0.410 |
| | short term | -0.405 | 0.338 | 684.834 | 1.000 | -1.401 | 0.592 |
| | various | -3.676* | 0.255 | 593.025 | 0.000 | -4.427 | -2.926 |
| timing | beta blockers | 1.661* | 0.227 | 305.975 | 0.000 | 0.990 | 2.332 |
| | antidepressants | 1.435* | 0.296 | 367.525 | 0.000 | 0.561 | 2.309 |
| | short term | 1.380* | 0.368 | 602.386 | 0.003 | 0.294 | 2.466 |
| | various | -1.892* | 0.293 | 504.046 | 0.000 | -2.757 | -1.026 |
| | no confounder | 1.785* | 0.207 | 331.538 | 0.000 | 1.172 | 2.398 |
| beta blockers | timing | -1.661* | 0.227 | 305.975 | 0.000 | -2.332 | -0.990 |
| | antidepressants | -0.226 | 0.273 | 357.043 | 1.000 | -1.033 | 0.582 |
| | short term | -0.281 | 0.351 | 623.628 | 1.000 | -1.314 | 0.752 |
| | various | -3.552* | 0.271 | 517.039 | 0.000 | -4.351 | -2.754 |
| | no confounder | 0.124 | 0.174 | 296.845 | 1.000 | -0.390 | 0.638 |
| antidepressants | timing | -1.435* | 0.296 | 367.525 | 0.000 | -2.309 | -0.561 |
| | beta blockers | 0.226 | 0.273 | 357.043 | 1.000 | -0.582 | 1.033 |
| | short term | -0.055 | 0.399 | 588.975 | 1.000 | -1.230 | 1.120 |
| | various | -3.327* | 0.331 | 506.835 | 0.000 | -4.302 | -2.351 |
| | no confounder | 0.350 | 0.257 | 384.985 | 1.000 | -0.410 | 1.110 |
| short term | timing | -1.380* | 0.368 | 602.386 | 0.003 | -2.466 | -0.294 |
| | beta blockers | 0.281 | 0.351 | 623.628 | 1.000 | -0.752 | 1.314 |
| | antidepressants | 0.055 | 0.399 | 588.975 | 1.000 | -1.120 | 1.230 |
| | various | -3.272* | 0.397 | 713.863 | 0.000 | -4.441 | -2.102 |
| | no confounder | 0.405 | 0.338 | 684.834 | 1.000 | -0.592 | 1.401 |
| various | timing | 1.892* | 0.293 | 504.046 | 0.000 | 1.026 | 2.757 |
| | beta blockers | 3.552* | 0.271 | 517.039 | 0.000 | 2.754 | 4.351 |
| | antidepressants | 3.327* | 0.331 | 506.835 | 0.000 | 2.351 | 4.302 |
| | short term | 3.272* | 0.397 | 713.863 | 0.000 | 2.102 | 4.441 |
| | no confounder | 3.676* | 0.255 | 593.025 | 0.000 | 2.926 | 4.427 |

##### 2.4.4 Results model B (Ikelos-RS)

**Table 11: Results mixed model B "Ikelos-RS" (I)**

| Tests of Fixed Effects | | | | |
|---|---|---|---|---|
| Source | Num. df | Den. Df | F | Sig. |
| Intercept | 1 | 871.855 | 14.010 | 0.000 |
| GROUP | 1 | 871.855 | 0.580 | 0.446 |
| ln_TIME | 1 | 842.708 | 4.811 | 0.029 |
| ln_TIME* ln_TIME | 1 | 812.591 | 3.315 | 0.069 |
| GROUP* ln_TIME | 1 | 842.708 | 1.029 | 0.311 |
| GROUP* ln_TIME* ln_TIME | 1 | 812.591 | 1.303 | 0.254 |

| Estimates of Fixed Effects | | | | | | | |
|---|---|---|---|---|---|---|---|
| | | | | | | 95 % CI | |
| Parameter | Estimate | Std. Error | df | t | Sig. | Lower Bound | Upper Bound |
| Intercept | 3.685076 | 0.808585 | 895.680 | 4.557 | 0.000 | 2.098134 | 5.272018 |
| GROUP (reference: continuous intake) | | | | | | | |
| GROUP (stopped after 6 months) | 1.883180 | 2.472186 | 871.855 | 0.762 | 0.446 | -2.968952 | 6.735312 |
| ln_TIME | -0.890177 | 0.505405 | 889.255 | -1.761 | 0.079 | -1.882103 | 0.101749 |
| ln_TIME* ln_TIME | 0.075183 | 0.076840 | 875.419 | 0.978 | 0.328 | -0.075630 | 0.225996 |
| GROUP* ln_TIME (reference: continuous intake ) | | | | | | | |
| GROUP ( stopped after 6 months) * ln_TIME | -1.532001 | 1.510195 | 842.708 | -1.014 | 0.311 | -4.496186 | 1.432184 |
| GROUP* ln_TIME* ln_TIME (reference: continuous intake ) | | | | | | | |
| GROUP ( stopped after 6 months) * ln_TIME* ln_TIME | 0.252735 | 0.221406 | 812.591 | 1.142 | 0.254 | -0.181859 | 0.687329 |

| Estimates (6 months) | | | | | |
|---|---|---|---|---|---|
| | | | | 95 % CI | |
| GROUP | Mean | Std. Error | df | Lower Bound | Upper Bound |
| continuous intake | 2.235 | 0.142 | 756.857 | 1.956 | 2.514 |
| stopped after 6 months | 2.092 | 0.432 | 848.544 | 1.244 | 2.940 |

| Pairwise Comparisons (6 months) | | | | | | | |
|---|---|---|---|---|---|---|---|
| | | | | | | 95 % CI | |
| (I)GROUP | (J)GROUP | Mean Difference (I-J) | Std. Error | df | Sig. | Lower Bound | Upper Bound |
| continuous intake | stopped after 6 months | 0.143 | 0.455 | 841.156 | 0.753 | -0.749 | 1.036 |

| Estimates (1 year) | | | | | |
|---|---|---|---|---|---|
| | | | | 95 % CI | |
| GROUP | Mean | Std. Error | df | Lower Bound | Upper Bound |
| continuous intake | 1.899 | 0.082 | 227.832 | 1.737 | 2.061 |
| stopped after 6 months | 1.517 | 0.246 | 432.607 | 1.034 | 1.999 |

| Pairwise Comparisons (1 year) | | | | | | | |
|---|---|---|---|---|---|---|---|
| | | | | | | 95 % CI | |
| (I)GROUP | (J)GROUP | Mean Difference (I-J) | Std. Error | df | Sig. | Lower Bound | Upper Bound |
| continuous intake | stopped after 6 months | 0.382 | 0.259 | 404.135 | 0.141 | -0.127 | 0.892 |

| Estimates (2 years) | | | | | |
|---|---|---|---|---|---|
| | | | | 95 % CI | |
| GROUP | Mean | Std. Error | df | Lower Bound | Upper Bound |
| continuous intake | 1.598 | 0.081 | 198.522 | 1.438 | 1.759 |
| stopped after 6 months | 1.169 | 0.233 | 323.687 | 0.710 | 1.628 |

| Pairwise Comparisons (2 years) | | | | | | | |
|---|---|---|---|---|---|---|---|
| | | | | | | 95 % CI | |
| (I)GROUP | (J)GROUP | Mean Difference (I-J) | Std. Error | df | Sig. | Lower Bound | Upper Bound |
| continuous intake | stopped after 6 months | 0.429 | 0.247 | 305.916 | 0.083 | -0.057 | 0.916 |

| Estimates (3 years) | | | | | |
|---|---|---|---|---|---|
| | | | | 95 % CI | |
| GROUP | Mean | Std. Error | df | Lower Bound | Upper Bound |
| continuous intake | 1.451 | 0.082 | 203.567 | 1.289 | 1.613 |
| stopped after 6 months | 1.098 | 0.218 | 317.257 | 0.669 | 1.527 |

| Pairwise Comparisons (3 years) | | | | | | | |
|---|---|---|---|---|---|---|---|
| | | | | | | 95 % CI | |
| (I)GROUP | (J)GROUP | Mean Difference (I-J) | Std. Error | df | Sig. | Lower Bound | Upper Bound |
| continuous intake | stopped after 6 months | 0.354 | 0.233 | 299.321 | 0.130 | -0.105 | 0.812 |

| Estimates (4 years) | | | | | |
|---|---|---|---|---|---|
| | | | | 95 % CI | |
| GROUP | Mean | Std. Error | df | Lower Bound | Upper Bound |
| continuous intake | 1.360 | 0.085 | 215.435 | 1.192 | 1.528 |
| stopped after 6 months | 1.108 | 0.193 | 277.028 | 0.728 | 1.488 |

| Pairwise Comparisons (4 years) | | | | | | | |
|---|---|---|---|---|---|---|---|
| | | | | | | 95 % CI | |
| (I)GROUP | (J)GROUP | Mean Difference (I-J) | Std. Error | df | Sig. | Lower Bound | Upper Bound |
| continuous intake | stopped after 6 months | 0.252 | 0.211 | 265.583 | 0.234 | -0.164 | 0.668 |

**Table 12: Results mixed model B "Ikelos-RS" (II)**

| Tests of Fixed Effects | | | | |
|---|---|---|---|---|
| Source | Num. df | Den. Df | F | Sig. |
| Intercept | 1 | 792.229 | 4.599 | 0.032 |
| GROUP | 3 | 834.503 | 2.378 | 0.068 |
| ln_TIME | 1 | 754.756 | 0.458 | 0.499 |
| ln_TIME* ln_TIME | 1 | 724.676 | 0.533 | 0.466 |
| GROUP* ln_TIME | 3 | 807.127 | 3.681 | 0.012 |
| GROUP* ln_TIME* ln_TIME | 3 | 782.145 | 3.572 | 0.014 |

| Estimates of Fixed Effects | | | | | | | |
|---|---|---|---|---|---|---|---|
| | | | | | | 95 % CI | |
| Parameter | Estimate | Std. Error | df | t | Sig. | Lower Bound | Upper Bound |
| Intercept | 3.645870 | 0.831837 | 962.319 | 4.383 | 0.000 | 2.013446 | 5.278293 |
| GROUP (reference: continuous intake) | | | | | | | |
| GROUP (stopped after 6 months) | 1.904393 | 2.548137 | 950.404 | 0.747 | 0.455 | -3.096231 | 6.905017 |
| GROUP (short term melatonin) | 8.465769 | 5.918606 | 738.174 | 1.430 | 0.153 | -3.153537 | 20.085075 |
| GROUP (various) | -8.655197 | 4.197571 | 841.354 | -2.062 | 0.040 | -16.894137 | -0.416258 |
| ln_TIME | -0.858659 | 0.520324 | 962.424 | -1.650 | 0.099 | -1.879760 | 0.162441 |
| ln_TIME* ln_TIME | 0.069371 | 0.079178 | 955.883 | 0.876 | 0.381 | -0.086012 | 0.224754 |
| GROUP* ln_TIME (reference: continuous intake ) | | | | | | | |
| GROUP ( stopped after 6 months) * ln_TIME | -1.555882 | 1.558431 | 925.901 | -0.998 | 0.318 | -4.614348 | 1.502583 |
| GROUP (short term melatonin) * ln_TIME | -5.696768 | 4.077561 | 709.146 | -1.397 | 0.163 | -13.702305 | 2.308769 |
| GROUP (various) * ln_TIME | 7.273292 | 2.633484 | 815.716 | 2.762 | 0.006 | 2.104089 | 12.442495 |
| GROUP* ln_TIME* ln_TIME (reference: continuous intake ) | | | | | | | |
| GROUP ( stopped after 6 months) * ln_TIME* ln_TIME | 0.257683 | 0.228698 | 897.706 | 1.127 | 0.260 | -0.191161 | 0.706527 |
| GROUP (short term melatonin) * ln_TIME* ln_TIME | 1.059169 | 0.663374 | 686.399 | 1.597 | 0.111 | -0.243317 | 2.361656 |
| GROUP (various) * ln_TIME* ln_TIME | -1.011053 | 0.398566 | 791.714 | -2.537 | 0.011 | -1.793425 | -0.228682 |

| Estimates (6 months) | | | | | |
|---|---|---|---|---|---|
| | | | | 95 % CI | |
| GROUP | Mean | Std. Error | df | Lower Bound | Upper Bound |
| continuous intake | 2.235 | 0.147 | 811.826 | 1.946 | 2.524 |
| stopped after 6 months | 2.086 | 0.446 | 923.373 | 1.211 | 2.960 |
| short term melatonin | 3.620 | 0.657 | 747.059 | 2.331 | 4.909 |
| various | 3.918 | 0.657 | 832.873 | 2.629 | 5.208 |

| Pairwise Comparisons (6 months) | | | | | | | |
|---|---|---|---|---|---|---|---|
| | | | | | | 95 % CI | |
| (I)GROUP | (J)GROUP | Mean Difference (I-J) | Std. Error | df | Sig. | Lower Bound | Upper Bound |
| continuous intake | stopped after 6 months | 0.150 | 0.469 | 914.936 | 1.000 | -1.091 | 1.391 |
| | short term melatonin | -1.385 | 0.673 | 750.218 | 0.240 | -3.164 | 0.395 |
| | various | -1.683 | 0.673 | 831.885 | 0.075 | -3.463 | 0.097 |
| stopped after 6 | short term melatonin | -1.534 | 0.794 | 810.737 | 0.321 | -3.633 | 0.564 |

| | | | | | | | |
|---|---|---|---|---|---|---|---|
| months | various | -1.833 | 0.794 | 865.338 | 0.127 | -3.932 | 0.266 |
| | continuous intake | -0.150 | 0.469 | 914.936 | 1.000 | -1.391 | 1.091 |
| short term melatonin | stopped after 6 months | 1.534 | 0.794 | 810.737 | 0.321 | -0.564 | 3.633 |
| | various | -0.299 | 0.929 | 790.927 | 1.000 | -2.755 | 2.158 |
| | continuous intake | 1.385 | 0.673 | 750.218 | 0.240 | -0.395 | 3.164 |
| various | stopped after 6 months | 1.833 | 0.794 | 865.338 | 0.127 | -0.266 | 3.932 |
| | short term melatonin | 0.299 | 0.929 | 790.927 | 1.000 | -2.158 | 2.755 |
| | continuous intake | 1.683 | 0.673 | 831.885 | 0.075 | -0.097 | 3.463 |

**Estimates (1 year)**

| GROUP | Mean | Std. Error | df | 95 % CI Lower Bound | 95 % CI Upper Bound |
|---|---|---|---|---|---|
| continuous intake | 1.902 | 0.087 | 233.462 | 1.732 | 2.073 |
| stopped after 6 months | 1.512 | 0.256 | 457.929 | 1.008 | 2.016 |
| short term melatonin | 2.726 | 0.453 | 486.406 | 1.835 | 3.616 |
| various | 5.241 | 0.312 | 512.245 | 4.629 | 5.853 |

**Pairwise Comparisons (1 year)**

| (I)GROUP | (J)GROUP | Mean Difference (I-J) | Std. Error | df | Sig. | 95 % CI Lower Bound | 95 % CI Upper Bound |
|---|---|---|---|---|---|---|---|
| continuous intake | stopped after 6 months | 0.390 | 0.271 | 425.578 | 0.903 | -0.328 | 1.108 |
| | short term melatonin | -0.823 | 0.462 | 473.246 | 0.450 | -2.046 | 0.399 |
| | various | -3.338* | 0.323 | 482.642 | 0.000 | -4.195 | -2.482 |
| stopped after 6 months | short term melatonin | -1.213 | 0.521 | 479.340 | 0.121 | -2.593 | 0.167 |
| | various | -3.728* | 0.404 | 489.582 | 0.000 | -4.797 | -2.659 |
| | continuous intake | -0.390 | 0.271 | 425.578 | 0.903 | -1.108 | 0.328 |
| short term melatonin | stopped after 6 months | 1.213 | 0.521 | 479.340 | 0.121 | -0.167 | 2.593 |
| | various | -2.515* | 0.550 | 494.557 | 0.000 | -3.972 | -1.058 |
| | continuous intake | 0.823 | 0.462 | 473.246 | 0.450 | -0.399 | 2.046 |
| various | stopped after 6 months | 3.728* | 0.404 | 489.582 | 0.000 | 2.659 | 4.797 |
| | short term melatonin | 2.515* | 0.550 | 494.557 | 0.000 | 1.058 | 3.972 |
| | continuous intake | 3.338* | 0.323 | 482.642 | 0.000 | 2.482 | 4.195 |

**Estimates (2 years)**

| GROUP | Mean | Std. Error | df | 95 % CI Lower Bound | 95 % CI Upper Bound |
|---|---|---|---|---|---|
| continuous intake | 1.600 | 0.086 | 206.349 | 1.431 | 1.770 |
| stopped after 6 months | 1.166 | 0.245 | 342.799 | 0.686 | 1.647 |
| short term melatonin | 2.704 | 0.517 | 546.902 | 1.689 | 3.720 |
| various | 5.882 | 0.305 | 453.081 | 5.283 | 6.481 |

**Pairwise Comparisons (2 years)**

| (I)GROUP | (J)GROUP | Mean Difference (I-J) | Std. Error | df | Sig. | 95 % CI Lower Bound | 95 % CI Upper Bound |
|---|---|---|---|---|---|---|---|
| continuous intake | stopped after 6 months | 0.434 | 0.259 | 322.944 | 0.571 | -0.254 | 1.122 |

| | | | | | | | |
|---|---|---|---|---|---|---|---|
| | short term melatonin | -1.104 | 0.524 | 531.864 | 0.214 | -2.492 | 0.284 |
| | various | -4.282* | 0.317 | 425.522 | 0.000 | -5.121 | -3.443 |
| stopped after 6 months | short term melatonin | -1.538* | 0.572 | 501.795 | 0.044 | -3.052 | -0.023 |
| | various | -4.716* | 0.391 | 405.643 | 0.000 | -5.751 | -3.680 |
| | continuous intake | -0.434 | 0.259 | 322.944 | 0.571 | -1.122 | 0.254 |
| short term melatonin | stopped after 6 months | 1.538* | 0.572 | 501.795 | 0.044 | 0.023 | 3.052 |
| | various | -3.178* | 0.600 | 521.088 | 0.000 | -4.767 | -1.589 |
| | continuous intake | 1.104 | 0.524 | 531.864 | 0.214 | -0.284 | 2.492 |
| various | stopped after 6 months | 4.716* | 0.391 | 405.643 | 0.000 | 3.680 | 5.751 |
| | short term melatonin | 3.178* | 0.600 | 521.088 | 0.000 | 1.589 | 4.767 |
| | continuous intake | 4.282* | 0.317 | 425.522 | 0.000 | 3.443 | 5.121 |

**Estimates (3 years)**

| GROUP | Mean | Std. Error | df | 95 % CI Lower Bound | 95 % CI Upper Bound |
|---|---|---|---|---|---|
| continuous intake | 1.450 | 0.087 | 211.582 | 1.279 | 1.621 |
| stopped after 6 months | 1.096 | 0.229 | 332.593 | 0.646 | 1.546 |
| short term melatonin | 3.154 | 0.437 | 524.585 | 2.295 | 4.013 |
| various | 5.875 | 0.277 | 429.231 | 5.330 | 6.421 |

**Pairwise Comparisons (3 years)**

| (I)GROUP | (J)GROUP | Mean Difference (I-J) | Std. Error | df | Sig. | 95 % CI Lower Bound | 95 % CI Upper Bound |
|---|---|---|---|---|---|---|---|
| continuous intake | stopped after 6 months | 0.354 | 0.245 | 313.158 | 0.891 | -0.295 | 1.004 |
| | short term melatonin | -1.704* | 0.446 | 505.732 | 0.001 | -2.884 | -0.523 |
| | various | -4.425* | 0.291 | 401.121 | 0.000 | -5.196 | -3.654 |
| stopped after 6 months | short term melatonin | -2.058* | 0.493 | 475.023 | 0.000 | -3.365 | -0.751 |
| | various | -4.779* | 0.360 | 386.633 | 0.000 | -5.733 | -3.826 |
| | continuous intake | -0.354 | 0.245 | 313.158 | 0.891 | -1.004 | 0.295 |
| short term melatonin | stopped after 6 months | 2.058* | 0.493 | 475.023 | 0.000 | 0.751 | 3.365 |
| | various | -2.722* | 0.518 | 495.227 | 0.000 | -4.093 | -1.350 |
| | continuous intake | 1.704* | 0.446 | 505.732 | 0.001 | 0.523 | 2.884 |
| various | stopped after 6 months | 4.779* | 0.360 | 386.633 | 0.000 | 3.826 | 5.733 |
| | short term melatonin | 2.722* | 0.518 | 495.227 | 0.000 | 1.350 | 4.093 |
| | continuous intake | 4.425* | 0.291 | 401.121 | 0.000 | 3.654 | 5.196 |

**Estimates (4 years)**

| GROUP | Mean | Std. Error | df | 95 % CI Lower Bound | 95 % CI Upper Bound |
|---|---|---|---|---|---|
| continuous intake | 1.355 | 0.090 | 225.368 | 1.179 | 1.532 |
| stopped after 6 months | 1.107 | 0.203 | 286.007 | 0.707 | 1.506 |
| short term melatonin | 3.688 | 0.446 | 492.331 | 2.812 | 4.564 |
| various | 5.694 | 0.254 | 369.907 | 5.195 | 6.193 |

**Pairwise Comparisons (4 years)**

| (I)GROUP | (J)GROUP | Mean Difference (I-J) | Std. Error | df | Sig. | 95 % CI | |
|---|---|---|---|---|---|---|---|
| | | | | | | Lower Bound | Upper Bound |
| continuous intake | stopped after 6 months | 0.249 | 0.222 | 274.736 | 1.000 | -0.341 | 0.839 |
| | short term melatonin | -2,333* | 0.455 | 476.645 | 0.000 | -3.538 | -1.128 |
| | various | -4,339* | 0.269 | 348.903 | 0.000 | -5.052 | -3.625 |
| stopped after 6 months | short term melatonin | -2,582* | 0.490 | 447.167 | 0.000 | -3.880 | -1.283 |
| | various | -4,587* | 0.325 | 333.916 | 0.000 | -5.450 | -3.725 |
| | continuous intake | -0.249 | 0.222 | 274.736 | 1.000 | -0.839 | 0.341 |
| short term melatonin | stopped after 6 months | 2,582* | 0.490 | 447.167 | 0.000 | 1.283 | 3.880 |
| | various | -2,006* | 0.513 | 458.820 | 0.001 | -3.365 | -0.646 |
| | continuous intake | 2,333* | 0.455 | 476.645 | 0.000 | 1.128 | 3.538 |
| various | stopped after 6 months | 4,587* | 0.325 | 333.916 | 0.000 | 3.725 | 5.450 |
| | short term melatonin | 2,006* | 0.513 | 458.820 | 0.001 | 0.646 | 3.365 |
| | continuous intake | 4,339* | 0.269 | 348.903 | 0.000 | 3.625 | 5.052 |

* The mean difference is significant at the 0.05 level.

American Academy of Sleep Medicine. International Classification of Sleep Disorders. 3rd ed. Darien. IL: 2014.

American Psychiatric Association. Diagnostic and statistical manual of mental disorders (5th ed.). Arlington. VA: 2015.

Djang DSW, Janssen MJR, Bohnen N, *et al*. SNM practice guideline for dopamine transporter imaging with $^{123}$I-Ioflupane SPECT 1.0. *J Nucl Med* 2012;53:154−163.

Goetz CG. Tilley BC. Shaftman SR. et al. Movement Disorder Society-sponsored revision of the Unified Parkinson's Disease Rating-scale (MDS-UPDRS): scale presentation and clinimetric testing results. *Mov Disord* 2008;23:2129−2170.

Guy W. "Clinical Global Impressions". ECDEU Assessment Manual for Psychopharmacology - Revised. Rockville. MD: U.S. Department of Health. Education and Welfare; Public Health Service, Alcohol, Drug Abuse, and Mental Health Administration; National Institute of Mental Health; Psychopharmacology Research Branch; Division of Extramural Research Programs 1976;218−222.

Hummel T, Sekinger B, Wolf SR, Pauli E, Kobal G. 'Sniffin' Sticks': olfactory performance assessed by the combined testing of odour identification, odor discrimination and olfactory threshold. *Chem Senses* 1997;22:39−52.

Kunz D , Bes F. Melatonin effects in a patient with severe REM sleep behavior disorder: case report and theoretical considerations. *Neuropsychobiol* 1997;36:211−214.

Kunz D, Bes F. Melatonin as a therapy in REM sleep behavior disorder patients: an open-labeled pilot study on the possible influence of melatonin on REM-sleep regulation. *Mov Disord* 1999;14:507−511

Kunz D, Mahlberg R. A two-part, double-blind, placebo-controlled trial of exogenous melatonin in REM sleep behaviour disorder. *J Sleep Res* 2010;19:591−6.

Norman G. Likert scales, levels of measurement and the "laws" of statistics. *Advances in Health Sciences Education* 2010;15:625−32.

Schmid NS, Ehrensperger MM, Berres M, Beck IR, Monsch AU. The extension of the German CERAD-Neuropsychological Assessment Battery with tests assessing subcortical, executive and frontal functions improves accuracy in dementia diagnosis. *Geriatr Cogn Disord Extra* 2014;4:322−334.

Visser M, Marinus J, Stiggelbout AM, van Hilten JJ. Assessment of autonomic dysfunction in Parkinson's disease: the SCOPA-AUT. *Mov Disord* 2004;19:1306−1312.

Zubal IG, Early M, Yuan O, Jennings D, Marek K, Seibyl JP. Optimized, automated striatal uptake analysis applied to SPECT brain scans of Parkinson's Disease patients. *J Nucl Med*. 2007;48:857−864.
